# Global landscape of *Streptococcus pneumoniae* serotypes colonising healthy individuals worldwide before vaccine introduction; a systematic review and meta-analysis

**DOI:** 10.1101/2023.03.09.23287027

**Authors:** Samuel Clifford, Maria D Knoll, Katherine L O’Brien, Timothy M Pollington, Riya Moodley, David Prieto-Merino, RESPICAR Consortium, W John Edmunds, Stefan Flasche, Olivier le Polain de Waroux

## Abstract

**Background:** Monitoring pneumococcal carriage prevalence and serotype distribution is critical to understanding pneumococcal transmission dynamics and vaccine impact, particularly where routine disease surveillance is limited. This study aimed to describe and interpret heterogeneity in serotype-specific carriage globally before widespread use of pneumococcal conjugate vaccines (PCVs).

**Methods:** A systematic literature review was undertaken to summarise all pneumococcal carriage studies across continents and age groups before PCV introduction. Serotype distributions were assessed via Bayesian nested meta-regression and hierarchical clustering.

**Findings:** In total 237 studies from 74 countries were included, comprising 492 age-specific datasets that contained 47,769 serotyped isolates.The modelled carriage prevalence differed substantially across regions, ranging in <5y from 35% (95%CrI 34%-35%) in Europe to 69% (95%CrI 69-70%) in Africa. Serotypes 19F, 6B, 6A, 23F, and 14 were the five most prevalent in children <5 years. The modelled proportion of Synflorix-10 (PCV10) serotypes carried by <5y ranged from 45% (95% CrI: 44% to 46%) in Asia to 59% (58% to 60%) in Europe, and that of Prevenar-13 (PCV13) from 60% (59% to 61%) in Asia to 76% (75% to 77%) in Europe. The diversity of carried serotypes increased with age, and so did the prevalence of vaccine-type serotypes. However, variation in serotype distribution did not cluster by age, ethnicity, region, or overall carriage prevalence.

**Interpretation:** Globally, pre-PCV pneumococcal carriage was dominated by a few serotypes. Serotype distribution variability was not easily attributable to a single discriminatory factor.

**Funding:** The review was funded by a grant to OlPdW from the World Health Organisation (grant number: SPHQ14-APW-2639) and by a Fellowship to SF jointly funded by the Wellcome Trust and the Royal Society (grant number: 208812/Z/17/Z).

## Background

Ten- and thirteen-valent pneumococcal conjugate vaccines (PCVs) (namely Synflorix-10 and Prevnar-13, respectively) have now been introduced into most national childhood immunisation programmes,^1^ substantially reducing the burden of pneumococcal disease.^2–8^ The impact of PCVs is partly driven by the vaccine effectiveness against disease among vaccinated persons but also by its impact against carriage.^9–11^ PCVs limit vaccine serotype acquisition and density thereby reducing community transmission and inducing herd immunity,^12,13^ which drives a substantial part of the overall impact of PCV programmes.^14–19^ However, the magnitude of the vaccine impact depends, amongst others, on the prevalence and serotype distribution of *Streptococcus pneumoniae* in carriage before vaccine introduction.^20^

Several alternative PCV formulations (Table S1) have been under development.^21^ Pneumosil-10 recently received WHO pre-qualification^22,23^ and 15- and 20-valent PCVs have been licensed by the US FDA for use in adults, and in June 2022 the 15-valent PCV has been recommended as an option in children by the US CDC’s Advisory Committee on Immunization Practices (ACIP).^24,25^ Studying pneumococcal carriage not only supports disease surveillance in countries with limited surveillance capacity for invasive pneumococcal disease (IPD),^26,27^ but the heterogeneity of pneumococcal carriage globally^28^ can be a good proxy for monitoring population-level vaccine impact.

Although a review of disease surveillance highlighted global geographic similarities and differences in IPD,^29^ little is known about the characteristics of *S. pneumoniae* serotype distribution in carriage, both intra- and internationally. We therefore conducted a landscape systematic **R**eview of the global **E**pidemiology of ***S****treptococcus* ***p****neumoniae* **I**n naso/oropharyngeal **Car**riage (RESPICAR) to provide an exhaustive overview before PCV introduction, and investigate drivers of heterogeneity in the distribution of carried serotypes.

## Methods

### Data

#### Search

Studies published before 1 January 2019 were identified using any combination of search terms in the groups “Pathogen” and “Endpoint” (Appendix 1 “Search terms”) (Figure S1). Identified articles were de-duplicated automatically initially, based on title, and further manually de-duplicated.

#### Data screening

The screening was conducted in three phases; (i) title and abstract, (ii) full text, and (iii) selected studies were classified as primary (the study reports a full carriage study of *Streptococcus pneumoniae*), co-primary (the study is one of many papers reporting a subset of data from the same carriage study), or secondary studies (re-analysis of existing data) (Figure S2).

Studies were included if they met the following five criteria: (i) study providing information on *S. pneumoniae* in carriage, (ii) from nasopharyngeal and oropharyngeal swabs, (iii) taken either from individuals in the community or outpatients (iv) in individuals who had not been vaccinated with PCV, and (v) in a setting where PCV had not yet been introduced into routine immunisation programmes.

In phase 1 screening, studies not meeting at least one inclusion criteria were excluded. If insufficient information was provided in the abstract and/or title to exclude the study, or if the study met the criteria based on abstract alone, the reference proceeded to full text screening. Studies were then classified as primary, co-primary, or secondary studies (phase iii) and, where appropriate, grouped under one single study. Studies written in a language other than English were assessed separately, and translation tools such Google Translate and Babylon were used when researchers had no working experience of the language.

Additionally, we excluded studies in which all participants were included based on presence or absence of symptoms suggestive of pneumococcal-like illness (e.g. acute respiratory infection, sinusitis, acute otitis media, sepsis, meningitis, and pneumonia). This was to ensure the population for which carriage estimates were provided was as representative as possible of the general population with regards to asymptomatic pneumococcal carriage. We also excluded conference abstracts with data that was later published as a paper. Finally, we excluded studies in which no sero-grouping or serotyping of the specimens was done (e.g. carriage prevalence only), as well as studies published before 1990 – a cut-off to limit the impact of changing demography and pneumococcal detection methods which only became standardised by a WHO working group in the early 2000s.^30^

For PCV trials we included data from control arms of either cluster randomised trials (under the assumption that there would be minimal spill-over effect from vaccinated clusters), as well as individually randomised PCV trials in which <20% of the study population in the targeted age group had received a PCV; 20% coverage of the entire population in an individual trial was deemed low enough to limit the indirect impact of vaccination.

Studies on groups particularly vulnerable to pneumococcal disease (such as patients with HIV or sickle cell disease) were recorded but excluded in this analysis.

For studies that met the eligibility criteria, but whose results on serotype and/or serogroup, or other data elements, were not directly or completely available from the paper we contacted authors and invited them to contribute to the RESPICAR Consortium with more detailed data.

#### Data entry

Data were extracted independently and entered into predefined templates in the DistillerSR software platform,^31^ by two independent researchers, with a third researcher resolving data entry conflicts. Data extracted included (i) the study design, (ii) the laboratory characteristics (including sample collection methods, culture methods and methods for serotyping), and (iii) the outcomes, including carriage prevalence, serotype/group distribution, year(s) and country the study was conducted, health status of the study population, and summary statistics of age.

#### Assumptions

Data were collected from studies that used different designs, endpoints and sampling methodologies. Hence, a series of assumptions were made for this analysis (Appendix 2).

In longitudinal studies with individuals swabbed multiple times, we averaged out the numerator and denominator over the study period if the sampling interval between studies was shorter than the maximum time for serotype clearance to avoid capturing the same carriage event. We assumed this to be four months for children aged under five years and three months in older age groups.^32–34^ Events separated by longer durations were deemed as independent events.

Multiple serotype colonisation was infrequently reported; however, when it did occur, equal weights were given to those serotypes reported and the total numbers of serotyped pneumococcal samples were considered as the denominator for the analysis of serotype distribution.

For serogroups for which information on serotypes was missing, we only included the available information in the model, and assumed *a priori* that the distribution of serotypes within serogroups was flat (see Analyses).

For cross-reactive serotypes which were not further subtyped, namely 6A/C and 15B/C, we reallocated the estimated prevalence for the subgroup to the individual cross-reactive serotypes proportional to their relative prevalence after sampling from the Bayesian model and before calculating model summaries or performing cluster analysis.

We identified studies targeting ethnic minorities whose epidemiological profiles may be unrepresentative of the national population,^35^ due to remoteness of settlement (e.g. Pygmy peoples of Gabon, Cameroon, and Congo), different demographics, access to health care services, refugee status (e.g. occupants of a camp on the Thailand-Myanmar border), or being indigenous inhabitants of colonised land (e.g. Native Americans in the USA, First Nations people in Canada, Australian Aborigines, or Māori in New Zealand), as such population groups may have high IPD rates and higher carriage prevalence.^36^ This was then tested in analysis.

### Analyses

Studies were stratified for analyses according to World Bank Development Indicators region definitions (Africa, Asia, Europe, Americas, and Oceania)^37^ and age group: less than five years old (<5y, young children), aged between five and seventeen, inclusive, (5–17y, school-aged children), and 18 and over (18+y, adults). Additional variables were collected for clustering of studies, namely: ethnic minority status and overall carriage prevalence. PCV-type specific prevalence is recoverable through aggregating the prevalence of the vaccine types, and this is used to calculate potential coverage of carriage events by different vaccine formulations. For studies that spanned multiple age groups and did not report results in finer age strata we used the median age of the participants to assign an age group. Overall carriage prevalence was classified as low, moderate, or high, using global, age-stratified terciles of carriage.

We used a nested Bayesian modelling approach combining a multinomial model for serogroups with a multinomial model for serotypes within serogroups (see the model’s details in the appendix). This framework allowed inclusion of small or zero values for rarer serotypes, as well as providing a natural weighting of the contribution of each study. Posterior distributions were sampled through Markov Chain Monte Carlo (MCMC) methods. The Gini, Gini-Simpson and Inverse Simpson indices of diversity were calculated from posterior samples as measures of pneumococcal diversity for serotype distribution (Tables S2–S4 in Appendix 3).^38,39^ All analyses were conducted in R 4.2.0; further details about the analytical approach can be found in Appendix 2 along with a link to scripts and datasets.

Hierarchical clustering was performed, based on the Bhattacharyya distance between the pairs of observed serotype distributions in datasets with at least ten serotyped pneumococcal-positive samples.^40,41^ After clustering, the composition of each cluster was considered with regard to each of: age category, continent, ethnic minority status, and overall carriage prevalence.

### Role of the funding source

The funders had no role in the design of the study, nor the collection, analyses, or interpretation of data, nor the writing of the manuscript or the decision to publish.

## Results

### Included studies

The initial literature screening identified 29,101 studies of which a total of 237 studies published during 1990–2018 were eventually included in this study (Figure 1, Figures S3–S6 and Appendix 5). Together, these included 492 datasets for serotype and/or serogroup distributions across different age groups, with a total of 60,857 samples that tested positive for pneumococci. Serogroup was available for 56,173 (92%) samples and 47,769 (78%) were serotyped. Studies were conducted worldwide, across 74 countries, although regional coverage within each continent was moderate, with 53% of the serotyped samples coming from only nine countries (Israel: 5604, Kenya: 3827, The Gambia: 3669, US: 2956, Portugal: 2713, Greece: 1832, UK: 1675, Uganda: 1602, and The Netherlands: 1405), while some countries had only one study and reported as few as 14 positive samples (South Sudan). Cyprus and Bulgaria were included in a multi-centre carriage study but no serotyped samples were reported in these countries.^42^ Most data were reported among young children (392 datasets) but 52 and 48 datasets reported carriage serotype distributions among school-aged children and adults, respectively (Table 1). As the systematic review only identified ten studies with a minimum age of at least 60 years, with fewer than 200 positive samples, we did not analyse young and elderly adults separately (see Figure S12 for a comparison of observed serotypes in the 18–59 and 60+ year old age groups).

**Table 1:** Overview of studies and samples found to be positive for the presence of pneumococci and serotype identified, in healthy individuals.

|  | Number of datasets (%) | Positive SPN<sup>1</sup> samples |  |  |
| --- | --- | --- | --- | --- |
|  |  | Total number (%) | Mean | IQR<sup>2</sup> |
| Age |  |  |  |  |
| Young children, <5y | 392 (80) | 41,268 (87) | 105 | [13–122] |
| School-aged, 5–17y | 52 (11) | 4,469 (9) | 86 | [17–131] |
| Adults, 18+ | 48 (10) | 2,032 (4) | 42 | [7–42] |
| Region |  |  |  |  |
| Africa | 83 (17) | 12,224 (26) | 147 | [12–153] |
| Americas | 105 (21) | 7,936 (17) | 76 | [12–93] |
| Asia | 128 (26) | 11,875 (25) | 93 | [7–100] |
| Europe | 167 (34) | 14,691 (31) | 88 | [12–116] |
| Oceania | 9 (2) | 1,043 (2) | 116 | [47–124] |
| Ethnic minority status |  |  |  |  |
| Minority<sup>3</sup> | 63 (13) | 3,404 (7) | 54 | [12–62] |
| Non minority | 429 (87) | 44,365 (93) | 103 | [12–121] |
| Total | 492 | 47,769 | 97 | [12–118] |

1: *Streptococcus pneumoniae*. 2: Interquartile range. 3: Minorities in Gabon and Venezuela, Native American in USA, Aboriginal Australians in Australia, Palestinian Arabs in Israel, and ethnic minorities.

**Figure 1:**
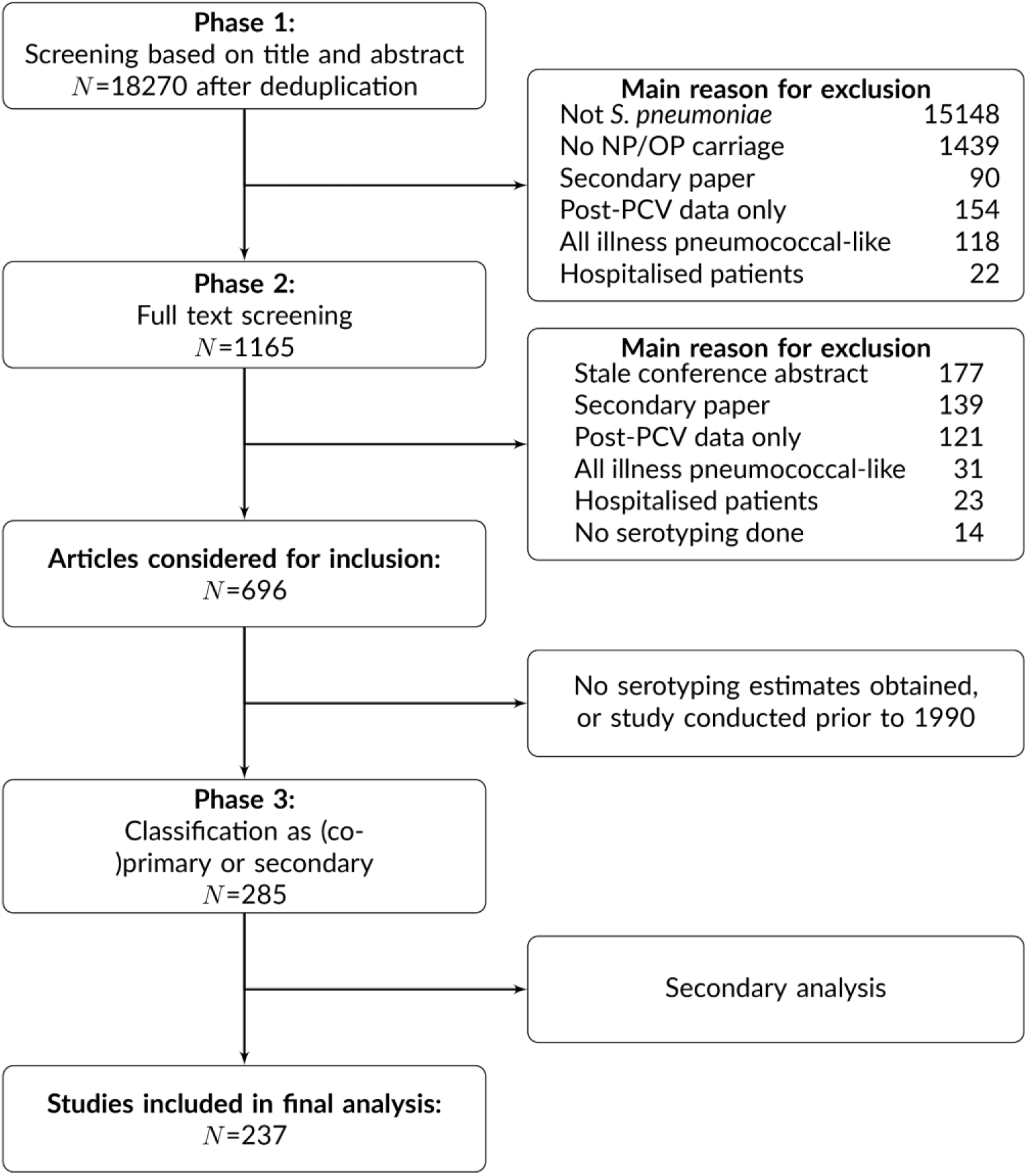
Flowchart of the screening process showing how many items remained at each step of screening and, where multiple reasons exist for exclusion, the primary reason for exclusion.

### Pneumococcal carriage

The overall modelled prevalence of pneumococcal carriage differed substantially across regions (Figure S7), ranging from 35% (95% Credible Interval: 34%, 35%) in Europe to 69% (69%, 70%) in Africa for young children, 18% (17%, 19%) in Asia to 73% (64%, 80%) in Oceania for school-aged children, and 5% (4%, 6%) in Europe to 20% (19%, 20%) in Africa among adults. In Oceania, where no adult data were available, the modelled prevalence was 19% (3%, 60%).

### Serotype distribution

Population-weighted averages of the proportion of carriage attributable to each serotype indicated that 10 serotypes (in decreasing order: 19F, 6B, 6A, 23F, 14, 19A, 15B, 9V, 11A, and 34) were responsible for 65% (64%, 66%) of paediatric carriage events (range: 63% (62%, 64%) in Asia to 76% (75%, 77%) in Europe). Carriage in children was generally dominated by a small number of serotypes, albeit with some variation across regions and age-groups (Figure 4). The Gini coefficient, indicating how diverse the modelled serotype distributions are in a given age and continent strata, ranged from a moderate 0·65 (0·59, 0·70) among (ethnic minority) 5–17 year olds in Oceania to a much less diverse distribution among <5 year olds in Europe where the Gini coefficient was 0·87 (0·86, 0·87).

### Vaccine serotype coverage

In young children, across the continents, the ten most prevalent serotypes always included serotypes: 6B, 23F, 19F, 6A, 14, 19A, 11A, and 15B. The diversity of carried serotypes in young children was similar across all regions (where the Gini index ranged from 0·78 (0·77, 0·78) in Asia to 0·82 (0·81, 0·82) in the Americas) except for Europe (being the least diverse, with a median Gini index of 0·86 (0·86, 0·87),) (Table S2). Also, the proportion of vaccine-type serotypes carried was relatively similar, with the proportion of carried serotypes (out of all serotypes) included in Synflorix-10 (PCV10) ranging from 45% (44% to 46%) in both Asia and Africa to 59% (58% to 59%) in Europe, and Prevnar-13 (PCV13) serotypes ranging from 60% (59% to 61%) in Asia to 76% (75% to 77%) in Europe (Figure S8).

Population-weighted global vaccine-type carriage in young children generally increased with valency, with Prevnar-20 including 72% (71% to 72%) of carriage serotypes, higher than Prevnar-13 and Vaxneuvance-15; 62% (62% to 63%) and 64% (64% to 65%), respectively. For the 10-valent vaccines, Pneumosil-10 included 59% (58%, 59%) and Synflorix-10 46% (46%, 47%) of carriage serotypes.

### Diversity in carried serotypes

For all continents the diversity of carried serotypes generally increased with age for all three indicators used (Gini, Gini-Simpson, and Inverse Simpson) and the proportion of vaccine preventable carriage episodes generally decreased with age (Table S2 and Figure 3). While in Europe and Asia the diversity of pneumococcal serotypes in healthy carriers was similar between young and school-age children in the other settings the diversity observed in school-age children was closer to that observed in adults (Table S2, Figures 4 and S9).

**Figure 2:**
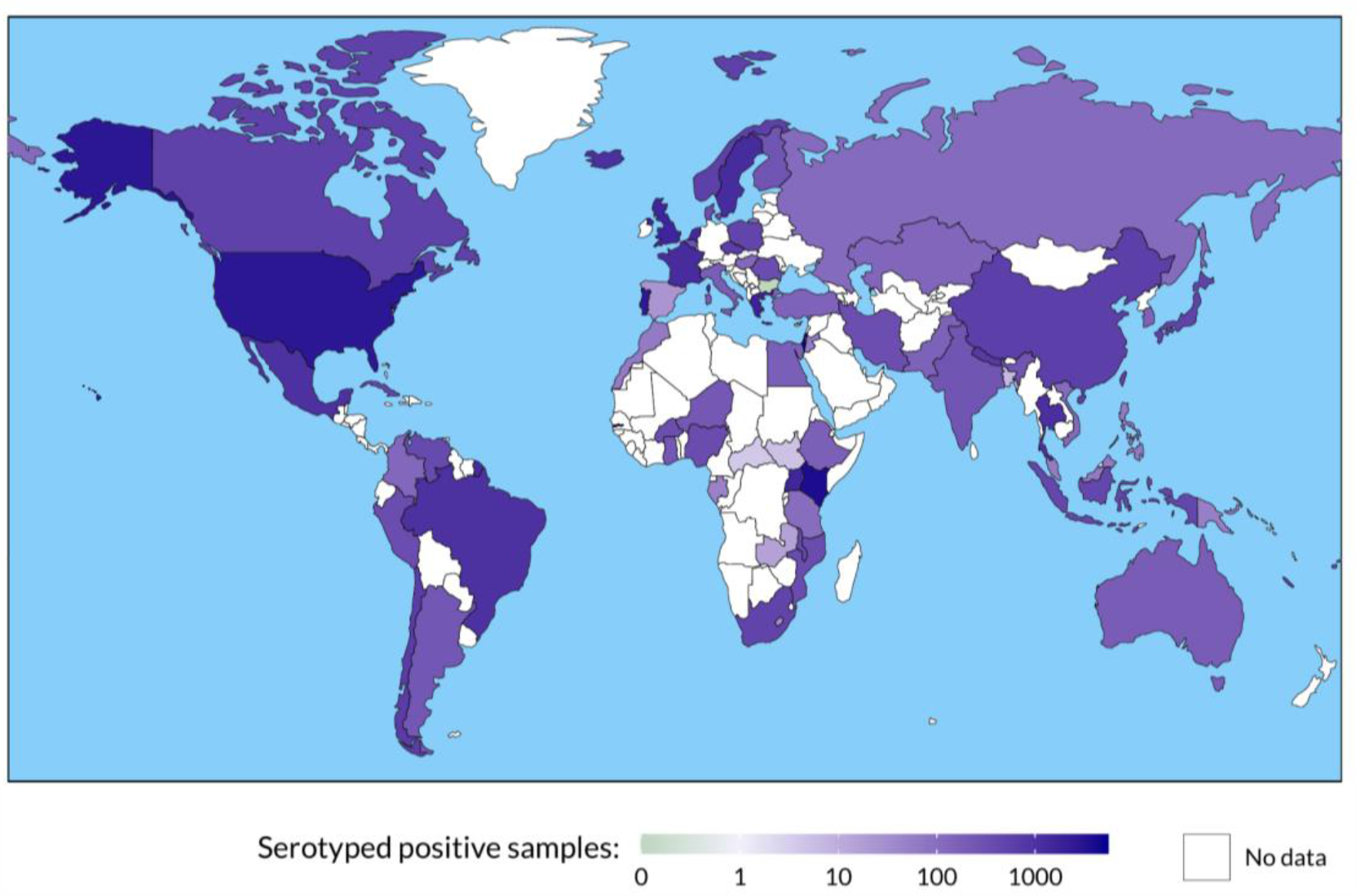
Map showing total number of serotyped isolates per country. Cream colouring indicates that no data was available for the respective country. Red colouring indicates data was available but no serotypes isolated. More than half of the serotyped isolates were collected in nine countries (Israel: 5604, Kenya: 3827, The Gambia: 3669, USA: 2956, Portugal: 2713, Greece: 1832, UK: 1675, Uganda: 1602, and The Netherlands: 1405). Map shapefile from Natural Earth (public domain).

**Figure 3:**
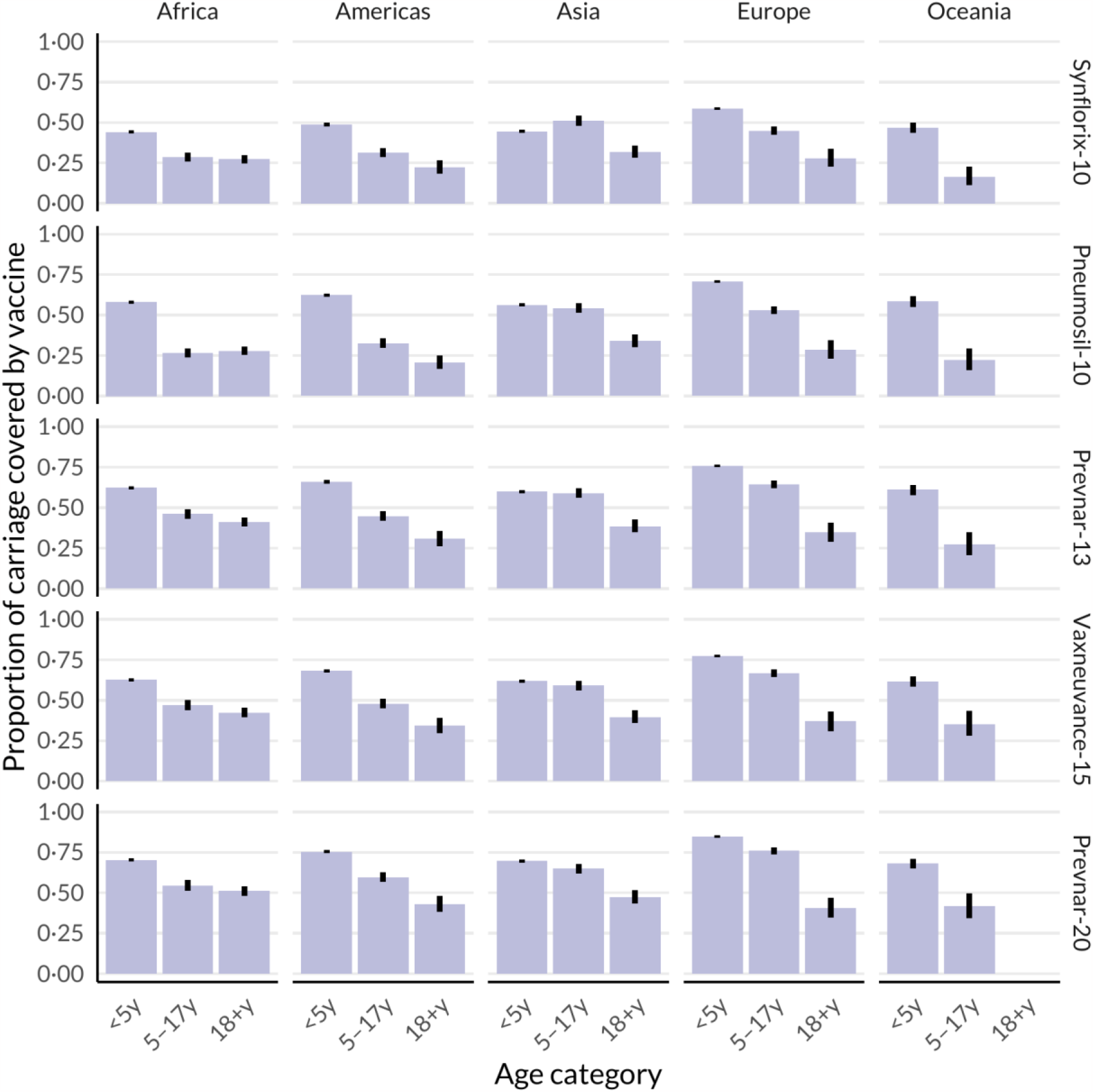
Proportion of carriage covered by the formulation in each vaccine product, stratified by continent and age group. Bars represent median estimates and error bars are 95% credible intervals.

### Predictors of serotype distribution

Initial cluster analysis indicated the presence of four clusters, with two clusters each containing a single dataset different enough to the others to warrant their own clusters (Figure S10). Assessment of the features not clustered on, namely region, age, indigenous status/ethnicity, or carriage prevalence, indicated that these alone could not be used to categorise datasets in distinct categories of serotype distribution (Figure 5). Of the 72 studies that contributed the 170 datasets to the clustering, 40 contributed only a single dataset. For the 32 studies contributing the remaining 130 datasets, each of 17 have all their datasets contained within one cluster, 12 have their datasets split across two clusters, two across three clusters, and one has their datasets spread across four clusters. This indicates that within-study variation may be greater than across-study variation, particularly for studies containing multiple age groups.

**Figure 4:**
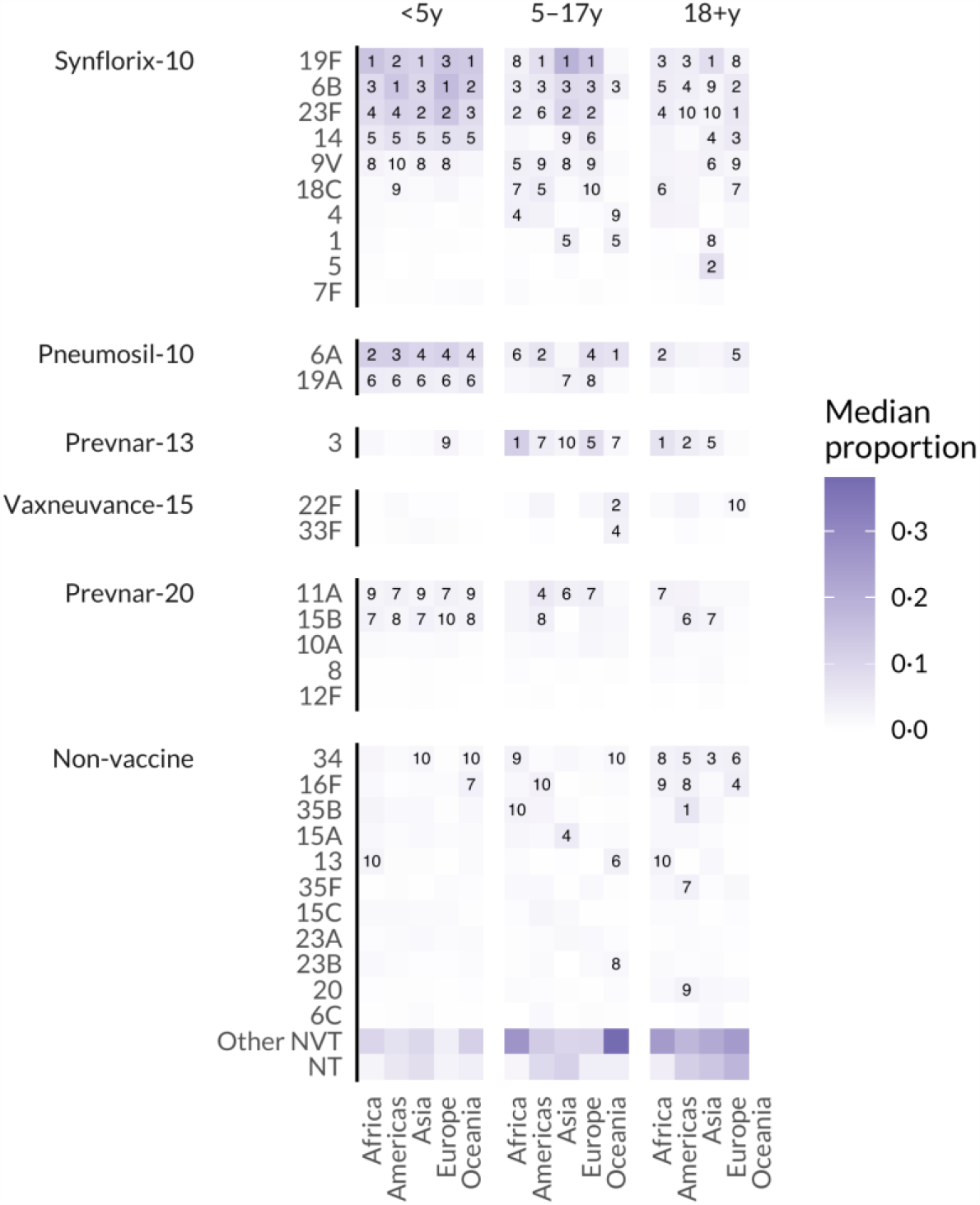
Median proportion of carriage attributed to each serotype within that age group and continent. The group labelled Synflorix-10 are the 10 serotypes included in that vaccine product; the group labelled Pneumosil-10 are the two serotypes found in that product in place of serotypes 4 and 18C in Synflorix-10. The serotypes in groups Prevnar-13, Vaxneuvance-15 and Prevnar-20 are those found in those products in addition to the products above. The non-vaccine types shown are the serotypes required to denote the 10 most carried serotypes in each age group in each continent (shown as numbers within grid cells). While serotypes 23B and 20 are more common in their respective age-continent settings than serotypes 15C and 23A, serotypes 15C and 23A are more common globally in this analysis and hence included in this graph. Serotype 6C is included due to its cross-reactivity with 6A. All additional serotypes contained in “Other NVT” are shown in Figure S9 and Appendix 6.

**Figure 5:**
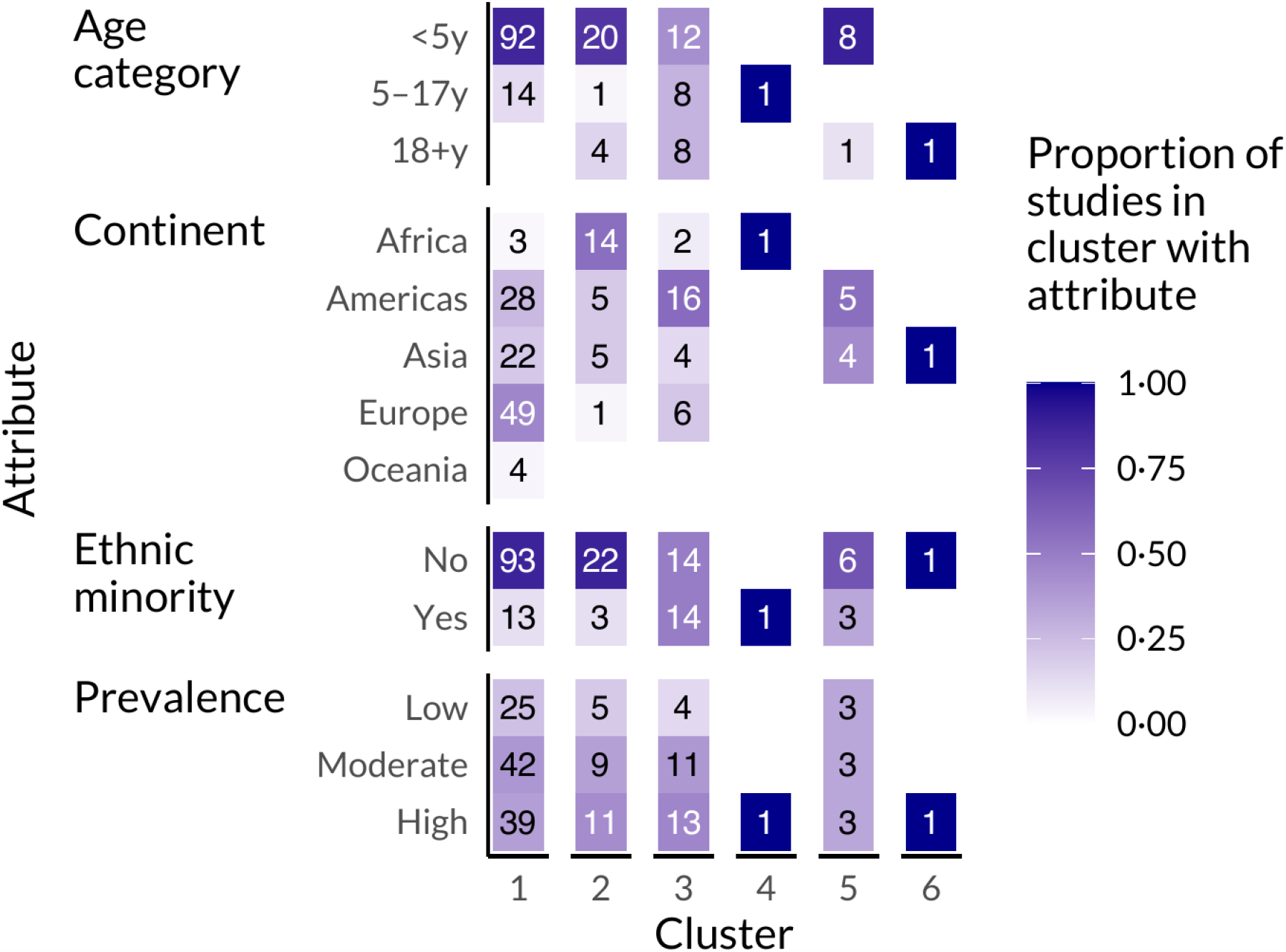
Values of variables not used to cluster in each of the six identified clusters (c.f. Figure S10). Cells are annotated with the number of studies in the cluster with the attribute at left and coloured by the same as a proportion. White text in the cell is for contrast purposes only.

### Studies among ethnic minority

After omitting low-power studies (less than 10 serotyped samples) the only ethnic minority population datasets included were two from Gabon, two from Venezuela, and 30 from the US; studies of Aboriginal Australians were excluded due to low power. The US was the only country to have studies included for both ethnic minority and general population groups in the cluster analysis (3 studies). As such, it is difficult to tell from the cluster analysis whether within-country variation is likely to be driven by ethnic minority status. Figure S11 shows which serotypes were observed (at least one measured carriage event) within each age group, by ethnic minority status, in each country where studies were conducted in ethnic minority groups and included in the cluster analysis.

## Discussion

Here we provide a comprehensive overview of the global serotype distribution among pneumococcal carriers of all ages in the pre-PCV era. We identified more than 200 studies reporting more than 60,000 samples that identified pneumococci. We find that, similar to IPD,^29^ the serotype distribution among pneumococcal carriers globally before PCV introduction was largely similar across settings and dominated by about 10 of the over 100 pneumococcal serotypes, although these 10 serotypes do not exactly correspond to any current 10-valent vaccine product, and carriage serotype distribution may not correspond to serotypes for diseases such as IPD, pneumonia, and otitis media. Clustering analyses showed that while serotype distribution may vary with age and continent, separation between the clusters cannot be ascribed to a simple partitioning on explanatory variables.

Pneumococcal carriage could be an important endpoint to potentially aid vaccine licensure^27^ and estimate the potential impact of PCVs in a given setting.^20,26,28^ We find that young and school-age children in Europe carried a proportionally higher number of vaccine serotypes, compared to other settings.^43^ This indicates a potentially larger vaccine impact but may also result in more pronounced serotype replacement, although ideally with less disease-prone serotypes.^44^ The serotype diversity among young children was highest in Asia (Gini 0·78, (0·77, 0·78), Table S2) and, unlike the other continents, a decrease in diversity was observed in the school-aged children. This is likely due to sample size as there are only 12 datasets across seven countries in the 5–17 year old group, compared to 102 datasets across 21 countries for the under-5s. Recent analysis of the distribution of serotypes in Nepal 2005–2013 shows greater diversity among carried types for the young than school-aged children.^45^

We found little evidence that geographic proximity implies a similar serotype distribution. This may stem from the nature of carriage studies, which, unlike invasive disease studies on population-based surveillance data, may target specific population groups which may not be a representative sample of the wider population in that country, as well as issues of small sample size. We estimate that in the Americas, the proportion of carriers that carry a vaccine serotype is lower than in Europe, Africa, or Asia. This result is largely driven by the serotype distribution among Native American populations (which comprise 73% of serotyped samples) where Prevnar-13 covers 52% (49%, 54%) of carriage events compared to 66% (63%, 70%) in the general population of the USA, and may contribute to the limited amount of replacement disease observed in the US.^46^ The difference in vaccine-type carriage is further borne out by Native American populations under 5 years carrying a more diverse range of serotypes (Gini 0·74 (0·72, 0·75)) than the general population of under 5s (Gini 0·80 (0·78, 0·82)). The list of carried serotypes in both ethnic minority and general populations in the USA is available in Figure S11.

Similarly, the school-aged participants in a study conducted among Babongo people in Gabon carried serotypes 15A, 3, 11A, 34, 17F, and 14 which were rarely observed elsewhere or even in other age groups within the same setting (where 6A, 7C, 10A, 13, 15B, and 19F are additionally carried by young children, but 3 and 17F are not, Figure S11).

Pneumococcal circulation may differ across populations where living environment and social contact patterns may drive transmission intensity within and between age groups, resulting in different carriage characteristics ^47^, as well as factors such as acquisition of antibiotic resistance and community antibiotic us prevalence. Other proxy factors should therefore be explored, such as demographic structure and contact patterns with those outside the studied cohort’s community, to try to better explain and disentangle factors associated with carriage diversity.

Capsule-specific acquired immunity is one of the main mechanisms balancing coexistence of pneumococcal serotypes.^48,49^ This implies that disproportionately high acquisition rates of dominant serotypes in early childhood eventually balances their fitness advantage by inducing capsule-specific immunity and permitting a more diverse set of pneumococci to colonise the host in subsequent years. In turn, this predicts that serotype diversity is correlated with transmission intensity and that an age shift in serotype diversity would happen at a younger age in settings with high carriage prevalence. Although our results corroborate this hypothesis to some extent, with serotype diversity generally increasing with age, and earlier in settings with a higher prevalence, more fully exploring this hypothesis is outside the scope of this manuscript.

## Limitations

The meta-analyses of multiple pneumococcal carriage studies faced a number of key challenges. These included the difference in sensitivity for detection of pneumococcal carriage (e.g. for culture vs PCR methods) and of serotypes as well as multiple carriage, the lack of serotyping beyond identification of the serogroup, particularly in older studies, and the longitudinal design of some studies. We used a hierarchical multinomial meta-regression framework which allows estimation of serogroup carriage prevalence where serotyping information was not provided. It also provides natural weighting and appropriate handling for the differentiation of instances where no carriage of a serotype was observed versus where the serotyping methods could not identify a serotype for all samples (e.g. limited amount of PCR primers). While in principle the analytical framework could be extendable to multiple carriage and multiple longitudinal observations, only a few studies used methods likely to yield high sensitivity for detecting multiple carriage^50^ and did not report results in sufficient detail to apply in this analysis. Thus we took a more pragmatic approach by averaging multiple observations in longitudinal designs and only included the dominant serotype if reported.

## Conclusion

In summary, we present an exhaustive overview of pneumococcal carriage studies globally before the introduction of PCVs. We report more than 60,000 positive pneumococcal samples from across the globe, including those excluded from our analysis due to comorbidities. There were, however, some large gaps that hindered the assessment of pneumococcal diversity particularly in populations with the likely highest pneumococcal disease burden, including large parts of Africa and crisis-affected populations. We found that a relatively small group of serotypes were predominantly carried across studies, although some differences existed that may, in part, determine differences in the impact of current and future pneumococcal vaccines. These differences could not be attributed to ethnic minority status, age group, or region.

## Data Availability

All data produced in the present study are available upon reasonable request to the authors

## Author contributions

SC - Model development, exploratory analysis, analysis, figures and tables, manuscript writing, data conflict resolution

MK - Project conception, manuscript writing

KO’B - Project conception, manuscript writing

TMP - search methodology, data extraction, manuscript editing RM - search methodology, data extraction, exploratory analysis

WJE - Projection conception, model development, manuscript writing

SF - Project conception, exploratory analysis, analysis, model development, manuscript writing, data conflict resolution

OlPdW - Project conception, search methodology, data extraction, exploratory analysis, analysis, model development, manuscript writing, data conflict resolution

Authors with access to underlying data: SC, SF, OlPdW RESPICAR Consortium: Interpretation, manuscript writing.

## Acknowledgements

The authors wish to acknowledge researchers and clinicians for their feedback or input on data from their papers including Jacobus de Waard, Helmia Farida, Didier Guillemot, Thomas Hennessey, Robin Hueben, Ioannis Katsarolis, Rezvan Moniri, Sabrina Moyo, Taketo Otsuka, Sarah Park, Maria C Rodriguez and Alexander Rowe

## Data sharing

Data and code to produce the analysis contained within this article, as well as a data dictionary, will be made available publicly with appropriate licence for reuse at date of publication. No individual participant data is used in the study and so none will be made available. The data is a collection of serotype-specific carriage and overall carriage, along with metadata about the study design and time and place in which the study was conducted. The sources of these datasets are available in Appendix 5. The study protocol is available in Appendix 4.

## Notes

### Competing Interest Statement

The authors have declared no competing interest.

### Author Declarations

This systematic review used published data.

