## Appendix 5 for "Global landscape of *Streptococcus pneumoniae* serotypes colonising healthy individuals worldwide before vaccine introduction; a systematic review and meta-analysis"

Samuel Clifford PhD[1] Maria D Knoll PhD[2] Katherine L O'Brien MD[2] Timothy M Pollington PhD[2] Riya Moodley MD[1]
RESPICAR Consortium (group) W John Edmunds PhD[1] Stefan Flasche PhD[1] Olivier le Polain de Waroux PhD[1]

[1] Department of Infectious Disease Epidemiology, London School of Hygiene & Tropical Medicine, London, WC1E 7HT, UK

[2] Department of International Health, Johns Hopkins Bloomberg School of Public Health, Baltimore, 21205, US

Table 1: Datasets included in the RESPICAR systematic review. Design: CS - Cross-sectional; Long. - Longitudinal; Unkn. - Unknown design. Sampling strategy: Rand. - Random/community; Outp. - Outpatients; Trial - Trial arm

| First author (date) | Reference | Year started | Country | ISO 3166-1 | Continent | Design | Sampling strategy | Ethnic Minority | Age group | Age range (years) | Median age (years) | # Positive | # Total | Prevalence |
|---|---|---|---|---|---|---|---|---|---|---|---|---|---|---|
| Adetifa (2012) | Pre-vaccination nasopharyngeal pneumococcal carriage in a Nigerian population: epidemiology and population biology. PLOS ONE 7 (1) | 2010 | Nigeria | 566 | Africa | CS | Rand. | No | <5y | 0–4 | Unknown | 375 | 524 | 72% |
| | | 2010 | Nigeria | 566 | Africa | CS | Rand. | No | 5–17y | 5–14 | Unknown | 63 | 120 | 52% |
| | | 2010 | Nigeria | 566 | Africa | CS | Rand. | No | 18+y | ≥18 | Unknown | 90 | 361 | 25% |
| Aho (2016) | Limited impact of neonatal or early infant schedules of 7-valent pneumococcal conjugate vaccination on nasopharyngeal carriage of Streptococcus pneumoniae in Papua New Guinean children: a randomized controlled trial. Vaccine Reports 6 | 2005 | Papua New Guinea | 598 | Oceania | Long. | Rand. | No | <5y | <1 | 0 | 62 | 79 | 78% |
| Al-Lahham (2009) | Resistance Patterns and Risk Factors of Streptococcus pneumoniae Carriage in Healthy Jordanian Children. Abstracts of the Interscience Conference on Antimicrobial Agents and Chemotherapy 49 (C2-1396) | 2009 | Jordan | 400 | Asia | CS | Outp. | No | <5y | 0–4 | Unknown | 65 | 118 | 55% |
| Altuzarra (2007) | Nasal carriage of Streptococcus pneumoniae in elderly subjects according to vaccination status. SpanishPortacion nasal de Streptococcus pneumoniae en adulto mayor y su respuesta frente a la vacunacion antineumococica. Revista Medica de Chile 135 (2) | 1998 | Chile | 152 | Americas | CS | Unkn. | No | 18+y | 60–60 | Unknown | 19 | 118 | 16% |
| Amaro (2018) | Prevalence of naso-pharyngeal Pneumococcal colonization in children between 1 to 5 years old in pre-school educative centers. 2014-2015. Medisur-Revista De Ciencias Medicas De Cienfuegos 16 (3) | 2014 | Cuba | 192 | Americas | CS | Unkn. | No | <5y | 1–5 | 3 | 356 | 1129 | 32% |
| Andrade (2010) | Survey of nonsusceptible nasopharyngeal streptococcus pneumoniae isolates in children attending day-care centers in Brazil. Pediatric Infectious Disease Journal 29 | 2005 | Brazil | 76 | Americas | CS | Unkn. | No | <5y | <1 | 0 | 6 | 20 | 30% |
| | | 2005 | Brazil | 76 | Americas | CS | Unkn. | No | <5y | 1–1 | 1 | 80 | 190 | 42% |
| | | 2005 | Brazil | 76 | Americas | CS | Unkn. | No | <5y | 2–4 | 3 | 244 | 979 | 25% |
| | | 2008 | Brazil | 76 | Americas | CS | Unkn. | No | <5y | <1 | 0 | 3 | 7 | 43% |

| | | | | | | | | | | | | | | |
|---|---|---|---|---|---|---|---|---|---|---|---|---|---|---|
| Andrade (2012) | Molecular epidemiological investigation to determine the source of a fatal case of serotype 22F pneumococcal meningitis. Journal of Medical Microbiology 61 | 2008 | Brazil | 76 | Americas | CS | Unkn. | No | <5y | 1–1 | 1 | 4 | 7 | 57% |
| | | 2008 | Brazil | 76 | Americas | CS | Unkn. | No | <5y | 2–4 | 3 | 61 | 102 | 60% |
| Appelbaum (1996) | Carriage of antibiotic-resistant Streptococcus pneumoniae by children in eastern and central Europe-a multicenter study with use of standardized methods. Clinical Infectious Diseases 23 | 1993 | Cyprus | 196 | Asia | CS | Outp. | No | <5y | 0–4 | Unknown | 12 | 100 | 12% |
| | | 1993 | Bulgaria | 100 | Europe | CS | Outp. | No | <5y | 0–4 | Unknown | 35 | 216 | 16% |
| | | 1993 | Poland | 616 | Europe | CS | Outp. | No | <5y | 0–4 | Unknown | 42 | 69 | 61% |
| | | 1993 | Romania | 642 | Europe | CS | Outp. | No | <5y | 0–4 | Unknown | 5 | 100 | 5% |
| | | 1993 | Russia | 643 | Europe | CS | Outp. | No | <5y | 0–4 | Unknown | 61 | 253 | 24% |
| | | 1993 | Slovakia | 703 | Europe | CS | Outp. | No | <5y | 0–4 | Unknown | 103 | 216 | 48% |
| Arason (1996) | Do antimicrobials increase the carriage rate of penicillin resistant pneumococci in children? Cross sectional prevalence study. British Medical Journal 313 (7054) | 1992 | Iceland | 352 | Europe | CS | Outp. | No | <5y | 0–6 | Unknown | | | Unkn. |
| Aslan (2007) | Serotype distribution of Streptococcus pneumoniae strains in the nasopharynx of healthy Turkish children. Indian Journal of Medical Research 125 (4) | 2003 | Turkey | 792 | Asia | CS | Unkn. | No | 5–17y | 6–13 | 9 | 201 | 1440 | 14% |
| Auranen (2010) | Between-strain competition in acquisition and clearance of pneumococcal carriage–epidemiologic evidence from a longitudinal study of day-care children. American Journal of Epidemiology 171 | 1999 | Denmark | 208 | Europe | Long. | Unkn. | No | <5y | 0–3 | 2 | 423 | 571 | 74% |
| | | 1999 | Denmark | 208 | Europe | Long. | Unkn. | No | 18+y | ≥18 | Unknown | 35 | 161 | 22% |
| Badawy (2017) | Serotypes of Streptococcus pneumoniae in Egyptian children: are they covered by pneumococcal conjugate vaccines?. European Journal of Clinical Microbiology and Infectious Diseases 36 (12) | 2012 | Egypt | 818 | Africa | CS | Outp. | No | <5y | 0–5 | 2 | 62 | 200 | 31% |
| Bae (2013) | Prevalent Multidrug-resistant Nonvaccine Serotypes in Pneumococcal Carriage of Healthy Korean Children Associated with the Low Coverage of the Seven-valent Pneumococcal Conjugate Vaccine. Osong Public Health and Research Perspectives 4 | 2006 | South Korea | 410 | Asia | Long. | Unkn. | No | 5–17y | 6–10 | 8 | 155 | 1668 | 11% |
| Balicer (2010) | Control of Streptococcus pneumoniae serotype 5 epidemic of severe pneumonia among young army recruits by mass antibiotic treatment and vaccination. Vaccine 28 (34) | 2005 | Israel | 376 | Asia | CS | Unkn. | No | 18+y | 18–20 | 18 | 58 | 142 | 41% |
| Bayraktar (2005) | Nasopharyngeal carriage of Streptococcus pneumoniae in healthy children attending day care centres in Malatya. Infeksiyon Dergisi. Turkish Journal of Infection 19 (3) | 2003 | Turkey | 792 | Asia | CS | Unkn. | No | 5–17y | 7–13 | Unknown | 162 | 848 | 19% |
| Berezin (2012) | Pneumococcal nasopharyngeal carriage in infants of mothers immunized with 23V non-conjugate pneumococcal polysaccharide vaccine. Journal of Tropical Pediatrics 58 | 2005 | Brazil | 76 | Americas | CS | Outp. | No | <5y | <1 | Unknown | 31 | 139 | 22% |
| Berkovitch (2002) | Colonization rate of bacteria in the throat of healthy infants. International Journal of Pediatric Otorhinolaryngology 63 | 2000 | Israel | 376 | Asia | CS | Outp. | No | <5y | 0–1 | 1 | 23 | 1000 | 2% |
| Blossom (2007) | Characterization of penicillin intermediate serotypes of Streptococcus pneumoniae carried by human immunodeficiency virus-infected adults and healthy children in Uganda. Microbial Drug Resistance 13 | 2004 | Uganda | 800 | Africa | CS | Outp. | No | <5y | 0–2 | Unknown | 113 | | Unkn. |
| Bogaert (2001) | Pneumococcal carriage in children in The Netherlands: a molecular epidemiological study. Journal of Clinical Microbiology 39 | 1999 | Netherlands | 528 | Europe | CS | Rand. | No | <5y | 0–3 | 1 | 305 | | Unkn. |
| Bokaeian (2011) | Nasopharyngeal carriage, antibiotic resistance and serotype distribution of Streptococcus pneumoniae among healthy adolescents in Zahedan. Iranian Red Crescent Medical Journal 13 (5) | 2007 | Iran | 364 | Asia | CS | Unkn. | No | 5–17y | 10–19 | Unknown | 136 | 865 | 16% |
| Boken (1995) | Colonization with penicillin-resistant Streptococcus pneumoniae in a child-care center. Pediatric Infectious Disease Journal 14 (10) | 1994 | United States | 840 | Americas | CS | Unkn. | No | <5y | 1–2 | Unknown | 32 | 54 | 59% |
| Boken (1996) | Colonization with penicillin-nonsusceptible Streptococcus pneumoniae in urban and rural child-care centers. Pediatric Infectious Disease Journal 15 (8) | 1990 | United States | 840 | Americas | CS | Unkn. | No | <5y | 0–1 | 1 | 121 | 215 | 56% |
| Browall (2014) | Intraclonal variations among Streptococcus pneumoniae isolates influence the likelihood of invasive disease in children. Journal of Infectious Diseases 209 (3) | 1997 | Sweden | 752 | Europe | CS | Unkn. | No | <5y | <1 | Unknown | 55 | 550 | 10% |
| Brueggemann (2003) | Clonal relationships between invasive and carriage Streptococcus pneumoniae and serotype- and clone-specific differences in invasive disease potential. Journal of Infectious Diseases 187 | 1994 | United Kingdom | 826 | Europe | Long. | Rand. | No | <5y | 0–4 | Unknown | | | Unkn. |
| Calderón-Jaimes (1993) | The resistance and serotyping of 83 strains of Streptococcus pneumoniae isolated from asymptomatic carriers and ill children. Boletin Medico del Hospital Infantil de Mexico 50 (12) | 1992 | Mexico | 484 | Americas | CS | Unkn. | No | <5y | 2–5 | Unknown | 35 | 44 | 80% |
| Camurdan (2008) | Nasopharyngeal carriage of Streptococcus pneumoniae in healthy Turkish infants. Journal of Infection 56 | 2003 | Turkey | 792 | Asia | CS | Outp. | No | <5y | 1–2 | 1 | 127 | 564 | 23% |

| | | | | | | | | | | | | | | |
|---|---|---|---|---|---|---|---|---|---|---|---|---|---|---|
| Cardozo (2006) | Antimicrobial resistance and serotypes of nasopharyngeal strains of Streptococcus pneumoniae in Brazilian adolescents. Microbial Drug Resistance 12 (1) | 2003 | Brazil | 76 | Americas | CS | Unkn. | No | 5–17y | 10–19 | Unknown | 83 | 1013 | 8% |
| Caugant (2006) | Carriage of Streptococcus pneumoniae in healthy Norwegian children attending day-care centres. European Journal of Clinical Microbiology and Infectious Diseases 25 | 2003 | Norway | 578 | Europe | Long. | Unkn. | No | <5y | 1–1 | 1 | 6 | 10 | 60% |
| | | 2003 | Norway | 578 | Europe | Long. | Unkn. | No | <5y | 1–1 | 1 | 27 | 46 | 59% |
| | | 2003 | Norway | 578 | Europe | Long. | Unkn. | No | <5y | 2–4 | Unknown | 5 | 8 | 62% |
| | | 2003 | Norway | 578 | Europe | Long. | Unkn. | No | <5y | 2–4 | Unknown | 9 | 23 | 39% |
| | | 2003 | Norway | 578 | Europe | Long. | Unkn. | No | 5–17y | 5–6 | Unknown | 6 | 16 | 38% |
| | | 2003 | Norway | 578 | Europe | Long. | Unkn. | No | 5–17y | 5–6 | Unknown | 7 | 24 | 29% |
| | | 2003 | Norway | 578 | Europe | Long. | Unkn. | No | <5y | 1–1 | 1 | 33 | 56 | 59% |
| | | 2003 | Norway | 578 | Europe | Long. | Unkn. | No | <5y | 2–4 | Unknown | 14 | 31 | 45% |
| | | 2003 | Norway | 578 | Europe | Long. | Unkn. | No | 5–17y | 5–6 | Unknown | 13 | 40 | 32% |
| | | 2004 | Norway | 578 | Europe | Long. | Unkn. | No | <5y | 1–1 | Unknown | 6 | 11 | 55% |
| | | 2004 | Norway | 578 | Europe | Long. | Unkn. | No | <5y | 2–4 | Unknown | 2 | 9 | 22% |
| | | 2004 | Norway | 578 | Europe | Long. | Unkn. | No | 5–17y | 5–6 | Unknown | 5 | 16 | 31% |
| Caugant (2010) | Impact of a pneumococcal conjugate vaccination program on carriage among children in Norway. Clinical and Vaccine Immunology 17 | 2006 | Norway | 578 | Europe | CS | Unkn. | No | <5y | <1 | Unknown | 2 | 3 | 67% |
| | | 2006 | Norway | 578 | Europe | CS | Unkn. | No | <5y | 1–1 | Unknown | 48 | 55 | 87% |
| | | 2006 | Norway | 578 | Europe | CS | Unkn. | No | <5y | 2–4 | Unknown | 325 | 409 | 79% |
| | | 2006 | Norway | 578 | Europe | CS | Unkn. | No | <5y | 5–5 | 5 | 74 | 106 | 70% |
| Cekmez (2009) | Pneumococcal serotypes recovered from health children and their possible association with risk factor in Istanbul, Turkey. International Journal of Biomedical Science 5 (2) | 2007 | Turkey | 792 | Asia | CS | Outp. | No | 5–17y | 4–6 | Unknown | 9 | 125 | 7% |
| Charveriat (2005) | Nasopharyngeal carriage of Streptococcus pneumoniae in healthy children, 2 to 24 months of age, in New-Caledonia. FrenchEtude du portage rhinopharynge de Streptococcus pneumoniae chez les enfants sains ages de 2 a 24 mois en Nouvelle-Caledonie. Medecine et Maladies Infectieuses 35 | 2002 | New Caledonia | 540 | Oceania | CS | Outp. | No | <5y | 0–1 | 1 | 544 | 1040 | 52% |
| Cherian (1999) | Nasopharyngeal colonization of infants in southern India with Streptococcus pneumoniae. Epidemiology and Infection 123 | 1994 | India | 356 | Asia | Long. | Outp. | No | <5y | 0–1 | 0 | 202 | 506 | 40% |
| Cheung (2009) | Nasopharyngeal carriage of Streptococcus pneumoniae in Gambian children who participated in a 9-valent pneumococcal conjugate vaccine trial and in their younger siblings. Pediatric Infectious Disease Journal 28 | 2003 | Gambia | 270 | Africa | CS | Trial | No | <5y | 0–1 | 1 | 996 | 1061 | 94% |
| | | 2003 | Gambia | 270 | Africa | CS | Trial | No | <5y | 1–3 | 1 | 910 | 961 | 95% |
| | | 2004 | Gambia | 270 | Africa | CS | Trial | No | <5y | <1 | 0 | 305 | 329 | 93% |
| Coles (2002) | Nasopharyngeal carriage of resistant pneumococci in young South Indian infants. Epidemiology and Infection 129 | 1998 | India | 356 | Asia | Long. | Rand. | No | <5y | <1 | 0 | 247 | 352 | 70% |
| | | 1998 | India | 356 | Asia | Long. | Rand. | No | <5y | <1 | 0 | 259 | 404 | 64% |
| | | 1998 | India | 356 | Asia | Long. | Rand. | No | <5y | <1 | 0 | 183 | 462 | 40% |
| Coles (2009) | Nasopharyngeal carriage of S-pneumoniae among young children in rural Nepal. Tropical Medicine and International Health 14 | 2003 | Nepal | 524 | Asia | CS | Rand. | No | <5y | <1 | 0 | 118 | 149 | 79% |
| | | 2003 | Nepal | 524 | Asia | CS | Rand. | No | <5y | 1–1 | 1 | 249 | 300 | 83% |
| | | 2003 | Nepal | 524 | Asia | CS | Rand. | No | <5y | 2–4 | 3 | 84 | 101 | 83% |
| Coles (2011) | Newborn vitamin A supplementation does not affect nasopharyngeal carriage of Streptococcus pneumoniae in Bangladeshi infants at age 3 months. Journal of Nutrition 141 | 2004 | Bangladesh | 50 | Asia | CS | Rand. | No | <5y | <1 | 0 | 354 | 500 | 71% |
| Conklin (2016) | High Streptococcus pneumoniae colonization prevalence among HIV-infected Kenyan parents in the year before pneumococcal conjugate vaccine introduction. BMC Infectious Diseases 16 | 2009 | Kenya | 404 | Africa | CS | Unkn. | No | 18+y | 30–40 | Unknown | 41 | 153 | 27% |
| da Gloria Carvalho (2013) | Non-pneumococcal mitis-group streptococci confound detection of pneumococcal capsular serotype-specific loci in upper respiratory tract. PeerJ 1 (e97) | 2009 | Kenya | 404 | Africa | CS | Outp. | No | <5y | 0–4 | Unknown | 202 | 237 | 85% |
| | | 2009 | Kenya | 404 | Africa | CS | Outp. | No | 18+y | ≥18 | Unknown | 5 | 40 | 12% |
| | | 2009 | Israel | 376 | Asia | CS | Outp. | Yes | <5y | <1 | Unknown | 23 | 84 | 27% |
| | | 2009 | Israel | 376 | Asia | CS | Outp. | Yes | <5y | 1–1 | Unknown | 29 | 83 | 35% |

| | | | | | | | | | | | | | | |
|---|---|---|---|---|---|---|---|---|---|---|---|---|---|---|
| Daana (2015) | Measuring the effects of pneumococcal conjugate vaccine (PCV7) on Streptococcus pneumoniae carriage and antibiotic resistance: The Palestinian-Israeli Collaborative Research (PICR). Vaccine 33 (8) | 2009 | Israel | 376 | Asia | CS | Outp. | Yes | <5y | 2–4 | Unknown | 49 | 180 | 27% |
| | | 2009 | Palestinian Territories | 275 | Asia | CS | Outp. | Yes | <5y | <1 | Unknown | 100 | 270 | 37% |
| | | 2009 | Palestinian Territories | 275 | Asia | CS | Outp. | Yes | <5y | 1–1 | Unknown | 65 | 165 | 39% |
| | | 2009 | Palestinian Territories | 275 | Asia | CS | Outp. | Yes | <5y | 2–4 | Unknown | 58 | 186 | 31% |
| Dagan (2005) | Serum serotype-specific pneumococcal anticapsular immunoglobulin G concentrations after immunization with a 9-valent conjugate pneumococcal vaccine correlate with nasopharyngeal acquisition of pneumococcus. Journal of Infectious Diseases 192 (3) | 1996 | Israel | 376 | Asia | Long. | Unkn. | No | <5y | 2–4 | 2 | 284 | 367 | 77% |
| Dagan (2010) | Nasopharyngeal Carriage of Streptococcus pneumoniae Shortly before Vaccination with a Pneumococcal Conjugate Vaccine Causes Serotype-Specific Hyporesponsiveness in Early Infancy. Journal of Infectious Diseases 201 | 2005 | Israel | 376 | Asia | CS | Trial | No | <5y | <1 | Unknown | 204 | 459 | 44% |
| Dagan (2012) | The effect of an alternative reduced-dose infant schedule and a second year catch-up schedule with 7-valent pneumococcal conjugate vaccine on pneumococcal carriage: a randomized controlled trial. Vaccine 30 | 2005 | Israel | 376 | Asia | Long. | Trial | No | <5y | <1 | Unknown | 33 | 95 | 35% |
| | | 2005 | Israel | 376 | Asia | Long. | Trial | No | <5y | <1 | Unknown | 35 | 97 | 36% |
| | | 2005 | Israel | 376 | Asia | Long. | Trial | No | <5y | <1 | 0 | 18 | 98 | 18% |
| | | 2005 | Israel | 376 | Asia | Long. | Trial | No | <5y | <1 | Unknown | 42 | 92 | 46% |
| | | 2005 | Israel | 376 | Asia | Long. | Trial | No | <5y | <1 | Unknown | 62 | 79 | 78% |
| | | 2005 | Israel | 376 | Asia | Long. | Trial | No | <5y | <1 | Unknown | 60 | 78 | 77% |
| | | 2005 | Israel | 376 | Asia | Long. | Trial | No | <5y | <1 | 0 | 53 | 87 | 61% |
| | | 2005 | Israel | 376 | Asia | Long. | Trial | No | <5y | <1 | Unknown | 56 | 79 | 71% |
| | | 2005 | Israel | 376 | Asia | Long. | Trial | No | <5y | 1–1 | 1 | 45 | 91 | 49% |
| | | 2005 | Israel | 376 | Asia | Long. | Trial | No | <5y | 1–1 | 1 | 55 | 78 | 71% |
| Darboe (2010) | The dynamics of nasopharyngeal streptococcus pneumoniae carriage among rural Gambian mother-infant pairs. BMC Infectious Diseases 10 | 2001 | Gambia | 270 | Africa | Long. | Rand. | No | <5y | <1 | Unknown | 169 | 196 | 86% |
| | | 2001 | Gambia | 270 | Africa | Long. | Rand. | No | <5y | 1–1 | 1 | 182 | 196 | 93% |
| | | 2001 | Gambia | 270 | Africa | Long. | Rand. | No | 18+y | ≥18 | Unknown | 43 | 196 | 22% |
| | | 2001 | Gambia | 270 | Africa | Long. | Rand. | No | 18+y | ≥18 | Unknown | 45 | 196 | 23% |
| de Lencastre (1999) | Carriage and antibiotic resistance of respiratory pathogens and molecular epidemiology of antibiotic-resistant Streptococcus pneumoniae colonizing children in day-care centers in Lisbon: The Portuguese day-care center initiative. Clinical Microbiology and Infection 5 | 1996 | Portugal | 620 | Europe | CS | Unkn. | No | <5y | <1 | 0 | 9 | 20 | 45% |
| | | 1996 | Portugal | 620 | Europe | CS | Unkn. | No | <5y | 1–1 | 1 | 34 | 46 | 74% |
| | | 1996 | Portugal | 620 | Europe | CS | Unkn. | No | <5y | 2–4 | Unknown | 147 | 303 | 49% |
| | | 1996 | Portugal | 620 | Europe | CS | Unkn. | No | 5–17y | 5–6 | 5 | 84 | 217 | 39% |
| | | 1997 | Portugal | 620 | Europe | CS | Unkn. | No | <5y | <1 | Unknown | 15 | 29 | 52% |
| | | 1997 | Portugal | 620 | Europe | CS | Unkn. | No | <5y | 1–1 | 1 | 42 | 65 | 65% |
| | | 1997 | Portugal | 620 | Europe | CS | Unkn. | No | <5y | 2–4 | Unknown | 200 | 414 | 48% |
| | | 1997 | Portugal | 620 | Europe | CS | Unkn. | No | 5–17y | 5–6 | 5 | 92 | 237 | 39% |
| | | 1998 | Portugal | 620 | Europe | CS | Unkn. | No | <5y | <1 | 0 | 22 | 31 | 71% |
| | | 1998 | Portugal | 620 | Europe | CS | Unkn. | No | <5y | 1–1 | 1 | 50 | 80 | 62% |
| | | 1998 | Portugal | 620 | Europe | CS | Unkn. | No | <5y | 2–4 | Unknown | 242 | 398 | 61% |
| | | 1998 | Portugal | 620 | Europe | CS | Unkn. | No | 5–17y | 5–6 | 5 | 139 | 271 | 51% |
| | | 1999 | Portugal | 620 | Europe | CS | Unkn. | No | <5y | <1 | Unknown | 26 | 43 | 60% |
| | | 1999 | Portugal | 620 | Europe | CS | Unkn. | No | <5y | 1–1 | Unknown | 74 | 109 | 68% |

| | | | | | | | | | | | | | | |
|---|---|---|---|---|---|---|---|---|---|---|---|---|---|---|
| de Lencastre (2005) | Trends in drug resistance, serotypes, and molecular types of Streptococcus pneumoniae colonizing preschool-age children attending day care centers in Lisbon, Portugal: A summary of 4 years of annual surveillance. Journal of Clinical Microbiology 43 | 1999 | Portugal | 620 | Europe | CS | Unkn. | No | <5y | 2–4 | Unknown | 335 | 528 | 63% |
| | | 1999 | Portugal | 620 | Europe | CS | Unkn. | No | 5–17y | 5–6 | 5 | 155 | 254 | 61% |
| de Miguel-Martinez (2008) | Efficacy of heptavalent pneumococcal conjugate vaccine in children with cochlear implant. Acta Otorrinolaringologica Espanola 59 | 2005 | Spain | 724 | Europe | CS | Unkn. | No | <5y | 2–5 | 3 | 15 | 60 | 25% |
| | | 2005 | Spain | 724 | Europe | CS | Unkn. | No | <5y | 2–5 | 3 | 15 | 60 | 25% |
| de Waard (2007) | Pneumococcal carriage among indigenous Warao children in Venezuela: serotypes, susceptibility patterns, and molecular epidemiology. Clinical Infectious Diseases 45 | 2000 | Venezuela | 862 | Americas | CS | Rand. | Yes | <5y | 1–2 | 2 | 72 | 140 | 51% |
| | | 2004 | Venezuela | 862 | Americas | CS | Rand. | Yes | <5y | <1 | 0 | 24 | 57 | 42% |
| | | 2004 | Venezuela | 862 | Americas | CS | Rand. | Yes | <5y | 3–4 | 3 | 51 | 115 | 44% |
| | | 2004 | Venezuela | 862 | Americas | CS | Rand. | Yes | <5y | 5–5 | 5 | 14 | 44 | 32% |
| de Waard (2010) | Pneumococcal carriage in mothers and children of the panare amerindians from the state of Bolivar, Venezuela. SpanishEstado de portador nasofaringeo de streptococcus pneumoniae en madres e hijos de la poblacion indigena panare del estado Bolivar, Venezuela. Revista Argentina de Microbiologia 42 | 2008 | Venezuela | 862 | Americas | CS | Rand. | Yes | <5y | <1 | 0 | 7 | 11 | 64% |
| | | 2008 | Venezuela | 862 | Americas | CS | Rand. | Yes | <5y | 1–2 | Unknown | 21 | 27 | 78% |
| | | 2008 | Venezuela | 862 | Americas | CS | Rand. | Yes | <5y | 3–4 | Unknown | 30 | 46 | 65% |
| | | 2008 | Venezuela | 862 | Americas | CS | Rand. | Yes | 18+y | 18–40 | Unknown | 7 | 64 | 11% |
| de Waard (2011) | Carriage and invasive isolates of Streptococcus pneumoniae in Caracas, Venezuela: The relative invasiveness of serotypes and vaccine coverage. European Journal of Clinical Microbiology and Infectious Diseases 30 | 2007 | Venezuela | 862 | Americas | CS | Outp. | No | <5y | 0–3 | Unknown | 182 | 894 | 20% |
| Dunais (2011) | A decade-long surveillance of nasopharyngeal colonisation with Streptococcus pneumoniae among children attending day-care centres in south-eastern France: 1999-2008. European Journal of Clinical Microbiology and Infectious Diseases 30 | 1999 | France | 250 | Europe | CS | Unkn. | No | <5y | <1 | 0 | 25 | 44 | 57% |
| | | 1999 | France | 250 | Europe | CS | Unkn. | No | <5y | 1–1 | 1 | 76 | 123 | 62% |
| | | 1999 | France | 250 | Europe | CS | Unkn. | No | <5y | 2–4 | Unknown | 60 | 131 | 46% |
| | | 2002 | France | 250 | Europe | CS | Unkn. | No | <5y | <1 | 0 | 33 | 57 | 58% |
| | | 2002 | France | 250 | Europe | CS | Unkn. | No | <5y | 1–1 | 1 | 67 | 115 | 58% |
| | | 2002 | France | 250 | Europe | CS | Unkn. | No | <5y | 2–4 | Unknown | 72 | 122 | 59% |
| Dunne (2018) | Carriage of streptococcus pneumoniae, haemophilus influenzae, moraxella catarrhalis, and staphylococcus aureus in Indonesian children: A cross-sectional study. PLOS ONE 13 (4) | 2016 | Indonesia | 360 | Asia | CS | Rand. | No | <5y | 1–2 | Unknown | 147 | 302 | 49% |
| Egere (2012) | Indirect effect of 7-valent pneumococcal conjugate vaccine on pneumococcal carriage in newborns in rural Gambia: a randomised controlled trial. PLOS ONE 7 | 2006 | Gambia | 270 | Africa | Long. | Rand. | No | <5y | 0–1 | Unknown | 572 | 963 | 59% |
| El-Nawawy (2015) | Nasopharyngeal Carriage, Capsular and Molecular Serotyping and Antimicrobial Susceptibility of Streptococcus pneumoniae among Asymptomatic Healthy Children in Egypt. Journal of Tropical Pediatrics 61 (1) | 2013 | Egypt | 818 | Africa | CS | Outp. | No | <5y | 0–5 | 1 | 175 | 600 | 29% |
| Espinosa-de los Monteros (2007) | Streptococcus pneumoniae isolates in healthy children attending day-care centers in 12 state in Mexico. Salud Publica de Mexico 49 | 2002 | Mexico | 484 | Americas | CS | Unkn. | No | <5y | 1–6 | Unknown | 829 | 2777 | 30% |
| Espinosa-de los Monteros (2010) | Serotype replacement of Streptococcus pneumoniae in children with pneumococcal conjugate vaccine 7V in Mexico. (Spanish). Salud Publica de Mexico 52 (1) | 2004 | Mexico | 484 | Americas | CS | Outp. | No | <5y | <1 | Unknown | 58 | 183 | 32% |
| Factor (2005) | Streptococcus pneumoniae and Haemophilus influenzae type B Carriage, Central Asia. Emerging Infectious Diseases 11 | 1997 | Kazakhstan | 398 | Asia | CS | Outp. | No | <5y | 0–4 | Unknown | 375 | 630 | 60% |
| Farida (2014) | Nasopharyngeal carriage of Streptococcus pneumonia in pneumonia-prone age groups in Semarang, Java Island, Indonesia. PLOS ONE 9 | 2010 | Indonesia | 360 | Asia | CS | Rand. | No | <5y | <1 | 0 | 21 | 45 | 47% |
| | | 2010 | Indonesia | 360 | Asia | CS | Rand. | No | <5y | 1–1 | Unknown | 24 | 57 | 42% |
| | | 2010 | Indonesia | 360 | Asia | CS | Rand. | No | <5y | 2–4 | Unknown | 60 | 141 | 43% |
| | | 2010 | Indonesia | 360 | Asia | CS | Rand. | No | 18+y | 45–70 | Unknown | 28 | 253 | 11% |
| Feikin (2003) | Antibiotic resistance and serotype distribution of Streptococcus pneumoniae colonizing rural Malawian children. Pediatric Infectious Disease Journal 22 | 1997 | Malawi | 454 | Africa | CS | Outp. | No | <5y | 0–4 | Unknown | 761 | 906 | 84% |
| | | 2000 | United Kingdom | 826 | Europe | Long. | Rand. | No | <5y | 2–5 | Unknown | 34 | 126 | 27% |
| | | 2000 | United Kingdom | 826 | Europe | Long. | Rand. | No | 18+y | 18–59 | Unknown | 6 | 138 | 4% |
| | | 2001 | United Kingdom | 826 | Europe | Long. | Rand. | No | <5y | 2–5 | Unknown | 5 | 203 | 2% |

| | | | | | | | | | | | | | | |
|---|---|---|---|---|---|---|---|---|---|---|---|---|---|---|
| Finn (2003) | Pneumococcal nasopharyngeal carriage in children following heptavalent pneumococcal conjugate vaccination in infancy. Archives of Disease in Childhood 88 | 2001 | United Kingdom | 826 | Europe | Long. | Rand. | No | <5y | 2–5 | Unknown | 78 | 188 | 41% |
| Frazão (2010) | Impact of a single dose of the 7-valent pneumococcal conjugate vaccine on colonization. Vaccine 28 (19) | 2001 | Portugal | 620 | Europe | Long. | Trial | No | <5y | 0–2 | Unknown | 68 | 85 | 80% |
| Galanis (2016) | Effects of PCV7 and PCV13 on invasive pneumococcal disease and carriage in Stockholm, Sweden. European Respiratory Journal 47 (4) | 2004 | Sweden | 752 | Europe | CS | Unkn. | No | <5y | 1–5 | Unknown | 260 | 719 | 36% |
| Ganaie (2018) | Impact of Hajj on the S. pneumoniae carriage among Indian pilgrims during 2016- a longitudinal molecular surveillance study. Travel Medicine and Infectious Disease 23 | 2016 | India | 356 | Asia | CS | Unkn. | No | 18+y | 18–100 | 56 | 145 | 807 | 18% |
| Gentile (2009) | First National Study of Prevalence for Nasopharyngeal Carriage (NPC) of Streptococcus pneumoniae (Spn) among Non-Vaccinated Children Attending Daycare Centers in Argentina. Abstracts of the Interscience Conference on Antimicrobial Agents and Chemotherapy 49 (C2-1397) | 2007 | Argentina | 32 | Americas | CS | Unkn. | No | <5y | 0–2 | Unknown | 211 | 381 | 55% |
| Gessner (2001) | Streptococcus pneumoniae nasopharyngeal carriage prevalence, serotype distribution, and resistance patterns among children on Lombok Island, Indonesia. Clinical Infectious Diseases 32 | 1997 | Indonesia | 360 | Asia | CS | Rand. | No | <5y | 0–1 | 1 | 221 | 484 | 46% |
| Gessner (2012) | Pneumococci in the African meningitis belt: meningitis incidence and carriage prevalence in children and adults. PLOS ONE 7 | 2008 | Burkina Faso | 854 | Africa | Long. | Rand. | No | <5y | <1 | Unknown | 43 | 62 | 69% |
| | | 2008 | Burkina Faso | 854 | Africa | Long. | Rand. | No | <5y | 1–4 | Unknown | 38 | 88 | 43% |
| | | 2008 | Burkina Faso | 854 | Africa | Long. | Rand. | No | 18+y | 5–39 | Unknown | 85 | 514 | 17% |
| Ghaffar (2002) | Effects of large dosages of amoxicillin/clavulanate or azithromycin on nasopharyngeal carriage of Streptococcus pneumoniae, Haemophilus influenzae, nonpneumococcal alpha -hemolytic streptococci, and Staphylococcus aureus in children with acute otitis media. Clinical Infectious Diseases 34 | 1997 | United States | 840 | Americas | CS | Rand. | No | <5y | 0–5 | Unknown | 5 | 45 | 11% |
| Giamarellou (2007) | Characterisation of macrolide-non-susceptible Streptococcus pneumoniae colonising children attending day-care centres in Athens, Greece during 2000 and 2003. Clinical Microbiology and Infection 13 | 2000 | Greece | 300 | Europe | CS | Unkn. | No | <5y | 1–6 | Unknown | 461 | 1451 | 32% |
| | | 2003 | Greece | 300 | Europe | CS | Unkn. | No | <5y | 1–6 | Unknown | 485 | 1396 | 35% |
| Giamarellou (2007) | Nationwide surveillance of Streptococcus pneumoniae in Greece: patterns of resistance and serotype epidemiology. International Journal of Antimicrobial Agents 30 | 2004 | Greece | 300 | Europe | CS | Unkn. | No | 5–17y | 0–6 | 5 | 793 | 2595 | 31% |
| Glennie (2013) | Defective pneumococcal-specific Th1 responses in HIV-infected adults precedes a loss of control of pneumococcal colonization. Clinical Infectious Diseases 56 | 2010 | Malawi | 454 | Africa | CS | Outp. | No | 18+y | 21–51 | 30 | 15 | 32 | 47% |
| Gómez-Barreto (2002) | Carriage of antibiotic-resistant pneumococci in a cohort of a daycare center. Salud Publica de Mexico 44 (1) | 1997 | Mexico | 484 | Americas | Long. | Unkn. | No | <5y | 1–5 | Unknown | 178 | 408 | 44% |
| Gordon (2003) | Poor potential coverage for 7-valent pneumococcal conjugate vaccine, Malawi. Emerging Infectious Diseases 9 (6) | 1998 | Malawi | 454 | Africa | CS | Rand. | No | 18+y | ≥18 | Unknown | 54 | 500 | 11% |
| Granat (2007) | Longitudinal study on pneumococcal carriage during the first year of life in Bangladesh. Pediatric Infectious Disease Journal 26 | 2000 | Bangladesh | 50 | Asia | Long. | Rand. | No | <5y | 0–1 | Unknown | 626 | 1459 | 43% |
| Greenberg (2011) | Nasopharyngeal carriage of individual Streptococcus pneumoniae serotypes during pediatric pneumonia as a means to estimate serotype disease potential. Pediatric Infectious Disease Journal 30 | 2001 | Israel | 376 | Asia | CS | Outp. | No | <5y | 0–4 | Unknown | 1504 | 2055 | 73% |
| Guillemot (1998) | Low dosage and long treatment duration of beta-lactam: Risk factors for carriage of penicillin-resistant Streptococcus pneumoniae. Journal of the American Medical Association 279 (5) | 1995 | France | 250 | Europe | CS | Unkn. | No | <5y | 3–6 | Unknown | | | Unkn. |
| | | 2000 | France | 250 | Europe | Long. | Unkn. | No | <5y | 2–4 | Unknown | 47 | 212 | 22% |
| | | 2000 | France | 250 | Europe | Long. | Unkn. | No | <5y | 2–4 | Unknown | 53 | 377 | 14% |
| | | 2000 | France | 250 | Europe | Long. | Unkn. | No | <5y | 2–4 | Unknown | 100 | 257 | 39% |
| | | 2000 | France | 250 | Europe | Long. | Unkn. | No | <5y | 2–4 | Unknown | 90 | 310 | 29% |
| | | 2000 | France | 250 | Europe | Long. | Unkn. | No | <5y | 2–4 | Unknown | 50 | 183 | 27% |
| | | 2000 | France | 250 | Europe | Long. | Unkn. | No | <5y | 2–4 | Unknown | 135 | 321 | 42% |
| | | 2000 | France | 250 | Europe | Long. | Unkn. | No | 5–17y | 5–6 | Unknown | 27 | 222 | 12% |
| | | 2000 | France | 250 | Europe | Long. | Unkn. | No | 5–17y | 5–6 | Unknown | 41 | 192 | 21% |
| | | 2000 | France | 250 | Europe | Long. | Unkn. | No | 5–17y | 5–6 | Unknown | 41 | 220 | 19% |
| | | 2000 | France | 250 | Europe | Long. | Unkn. | No | 5–17y | 5–6 | Unknown | 74 | 224 | 33% |

| | | | | | | | | | | | | | | |
|---|---|---|---|---|---|---|---|---|---|---|---|---|---|---|
| | | 2000 | France | 250 | Europe | Long. | Unkn. | No | 5–17y | 5–6 | Unknown | 46 | 172 | 27% |
| | | 2000 | France | 250 | Europe | Long. | Unkn. | No | 5–17y | 5–6 | Unknown | 114 | 278 | 41% |
| Hadinegoro (2016) | Nasopharyngeal Carriage of Streptococcus Pneumoniae in Healthy Children under Five Years Old in Central Lombok Regency, Indonesia. Southeast Asian Journal of Tropical Medicine and Public Health 47 (3) | 2012 | Indonesia | 360 | Asia | CS | Rand. | No | <5y | 0–5 | Unknown | 554 | 1200 | 46% |
| Hammitt (2014) | Population effect of 10-valent pneumococcal conjugate vaccine on nasopharyngeal carriage of Streptococcus pneumoniae and non-typeable Haemophilus influenzae in Kilifi, Kenya: findings from cross-sectional carriage studies. Lancet Global Health 2 | 2009 | Kenya | 404 | Africa | CS | Rand. | No | <5y | 0–4 | Unknown | 229 | 308 | 74% |
| Hanieh (2014) | Streptococcus pneumoniae carriage prevalence in Nepal: Evaluation of a method for delayed transport of samples from remote regions and implications for vaccine implementation. PLOS ONE 9 | 2009 | Nepal | 524 | Asia | CS | Outp. | No | <5y | 0–2 | Unknown | 337 | 574 | 57% |
| | | 2009 | Nepal | 524 | Asia | CS | Outp. | No | <5y | 0–2 | Unknown | 364 | 526 | 69% |
| Hanke (2016) | Bacterial density, serotype distribution and antibiotic resistance of pneumococcal strains from the nasopharynx of peruvian children before and after pneumococcal conjugate vaccine 7. Pediatric Infectious Disease Journal 35 (4) | 2009 | Peru | 604 | Americas | CS | Rand. | No | <5y | 0–3 | Unknown | 125 | | Unkn. |
| Harboe (2012) | A Pneumococcal carriage study in danish pre-school children before the introduction of pneumococcal conjugate vaccination. Open Microbiology Journal 6 | 1999 | Denmark | 208 | Europe | CS | Unkn. | No | <5y | 1–1 | Unknown | 247 | 437 | 57% |
| | | 1999 | Denmark | 208 | Europe | CS | Unkn. | No | <5y | 2–2 | Unknown | 247 | 437 | 57% |
| | | 1999 | Denmark | 208 | Europe | CS | Unkn. | No | <5y | 3–3 | Unknown | 247 | 437 | 57% |
| | | 1999 | Denmark | 208 | Europe | CS | Unkn. | No | <5y | 4–4 | Unknown | 247 | 437 | 57% |
| | | 1999 | Denmark | 208 | Europe | CS | Unkn. | No | <5y | 5–5 | Unknown | 247 | 437 | 57% |
| Heath (2018) | Nasopharyngeal carriage of streptococcus pneumoniae in children in Coastal Kenya. American Journal of Tropical Medicine and Hygiene 98 (4) | 2006 | Kenya | 404 | Africa | CS | Rand. | No | <5y | 0–1 | 0 | 65 | 323 | 20% |
| | | 2014 | Kenya | 404 | Africa | CS | Rand. | No | 5–17y | 4–7 | 6.1 | 65 | 323 | 20% |
| Hennessy (2002) | Changes in antibiotic-prescribing practices and carriage of penicillin-resistant Streptococcus pneumoniae: A controlled intervention trial in rural Alaska. Clinical Infectious Diseases 34 | 1998 | United States | 840 | Americas | CS | Rand. | Yes | <5y | <1 | 0 | 14 | 29 | 48% |
| | | 1998 | United States | 840 | Americas | CS | Rand. | Yes | <5y | 1–1 | Unknown | 19 | 34 | 56% |
| | | 1998 | United States | 840 | Americas | CS | Rand. | Yes | <5y | 2–4 | Unknown | 64 | 102 | 63% |
| | | 1998 | United States | 840 | Americas | CS | Rand. | Yes | 5–17y | 5–17 | Unknown | 201 | 428 | 47% |
| | | 1998 | United States | 840 | Americas | CS | Rand. | Yes | 18+y | ≥60 | Unknown | 6 | 62 | 10% |
| | | 1998 | United States | 840 | Americas | CS | Rand. | Yes | 18+y | 18–59 | Unknown | 61 | 448 | 14% |
| | | 1999 | United States | 840 | Americas | CS | Rand. | Yes | <5y | <1 | 0 | 13 | 25 | 52% |
| | | 1999 | United States | 840 | Americas | CS | Rand. | Yes | <5y | 1–1 | 1 | 13 | 25 | 52% |
| | | 1999 | United States | 840 | Americas | CS | Rand. | Yes | <5y | 2–4 | Unknown | 46 | 87 | 53% |
| | | 1999 | United States | 840 | Americas | CS | Rand. | Yes | 5–17y | 5–18 | Unknown | 170 | 425 | 40% |
| | | 1999 | United States | 840 | Americas | CS | Rand. | Yes | 18+y | ≥60 | Unknown | 5 | 50 | 10% |
| | | 1999 | United States | 840 | Americas | CS | Rand. | Yes | 18+y | 18–59 | Unknown | 48 | 400 | 12% |
| | | 2000 | United States | 840 | Americas | CS | Rand. | Yes | <5y | <1 | 0 | 13 | 19 | 68% |
| | | 2000 | United States | 840 | Americas | CS | Rand. | Yes | <5y | 1–1 | Unknown | 11 | 19 | 58% |
| | | 2000 | United States | 840 | Americas | CS | Rand. | Yes | <5y | 2–4 | Unknown | 34 | 81 | 42% |
| | | 2000 | United States | 840 | Americas | CS | Rand. | Yes | 5–17y | 5–17 | Unknown | 193 | 435 | 44% |
| | | 2000 | United States | 840 | Americas | CS | Rand. | Yes | 18+y | ≥60 | Unknown | 4 | 61 | 7% |
| | | 2000 | United States | 840 | Americas | CS | Rand. | Yes | 18+y | 18–59 | Unknown | 58 | 411 | 14% |
| | | 1998 | United States | 840 | Americas | CS | Rand. | Yes | <5y | <1 | 0 | 14 | 28 | 50% |
| | | 1998 | United States | 840 | Americas | CS | Rand. | Yes | <5y | 2–4 | Unknown | 40 | 86 | 47% |
| | | 1998 | United States | 840 | Americas | CS | Rand. | Yes | 5–17y | 5–17 | Unknown | 152 | 340 | 45% |

| | | | | | | | | | | | | | | |
|---|---|---|---|---|---|---|---|---|---|---|---|---|---|---|
| Hennessy (2002) | Effect of high-dose amoxicillin on the prevalence of penicillin-resistant Streptococcus pneumoniae in rural Alaska. Journal of the American Medical Association 287 | | | | | | | | | | | | | |
| | | 1998 | United States | 840 | Americas | CS | Rand. | Yes | 18+y | ≥60 | Unknown | 9 | 53 | 17% |
| | | 1998 | United States | 840 | Americas | CS | Rand. | Yes | 18+y | 18–59 | Unknown | 49 | 357 | 14% |
| Henriques Normark (2003) | Clonal analysis of Streptococcus pneumoniae nonsusceptible to penicillin at day-care centers with index cases, in a region with low incidence of resistance: emergence of an invasive type 35B clone among carriers. Microbial Drug Resistance 9 (4) | 1997 | Sweden | 752 | Europe | CS | Unkn. | No | <5y | 1–6 | 3 | 246 | 611 | 40% |
| Hermans (2011) | Epidemiology of Streptococcus pneumoniae and Staphylococcus aureus colonization in healthy Venezuelan children. European Journal of Clinical Microbiology and Infectious Diseases 30 | 2007 | Venezuela | 862 | Americas | CS | Unkn. | No | <5y | 2–5 | 5 | 71 | 250 | 28% |
| Hill (2008) | Nasopharyngeal carriage of Streptococcus pneumoniae in Gambian infants: A longitudinal study. Clinical Infectious Diseases 46 | 2005 | Gambia | 270 | Africa | Long. | Trial | No | <5y | 0–1 | 0 | 2710 | 3145 | 86% |
| Hjalmarsdottir (2015) | Prevalence of pilus genes in pneumococci isolated from healthy preschool children in Iceland: association with vaccine serotypes and antibiotic resistance. Journal of Antimicrobial Chemotherapy 70 (8) | 2009 | Iceland | 352 | Europe | CS | Unkn. | No | <5y | 1.2–6.3 | Unknown | 372 | 516 | 72% |
| Hjalmarsdottir (2016) | Cocolonization of Pneumococcal Serotypes in Healthy Children Attending Day Care Centers: Molecular Versus Conventional Methods. Pediatric Infectious Disease Journal 35 (5) | 2009 | Iceland | 352 | Europe | CS | Unkn. | No | <5y | 1.2–6.3 | 4.2 | 371 | 514 | 72% |
| Ho (2004) | Serotype distribution and antimicrobial resistance patterns of nasopharyngeal and invasive Streptococcus pneumoniae isolates in Hong Kong children. Vaccine 22 | 1999 | Hong Kong SAR China | 344 | Asia | CS | Unkn. | No | <5y | 2–4 | 3 | 209 | 1079 | 19% |
| | | 1999 | Hong Kong SAR China | 344 | Asia | CS | Unkn. | No | 5–17y | 5–6 | Unknown | 174 | 898 | 19% |
| Hogberg (2007) | Age- and serogroup-related differences in observed durations of nasopharyngeal carriage of penicillin-resistant pneumococci. Journal of Clinical Microbiology 45 | 1995 | Sweden | 752 | Europe | CS | Unkn. | No | <5y | 1–1 | 1 | | | Unkn. |
| | | 1995 | Sweden | 752 | Europe | CS | Unkn. | No | 5–17y | 5–6 | Unknown | 339 | | Unkn. |
| | | 1995 | Sweden | 752 | Europe | CS | Unkn. | No | 5–17y | 7–18 | Unknown | | | Unkn. |
| | | 1995 | Sweden | 752 | Europe | CS | Unkn. | No | 18+y | ≥18 | Unknown | 256 | | Unkn. |
| | | 1998 | Sweden | 752 | Europe | CS | Unkn. | No | <5y | 3–4 | Unknown | 632 | | Unkn. |
| Hu (2016) | Streptococcus pneumoniae and Haemophilus influenzae type b carriage in Chinese children aged 12-18 months in Shanghai, China: a cross-sectional study. BMC Infectious Diseases 16 | 2009 | China | 156 | Asia | CS | Outp. | No | <5y | 1–1.5 | Unknown | 102 | 614 | 17% |
| Huebner (1997) | Nasopharyngeal carriage of community-acquired, antibiotic-resistant Streptococcus pneumoniae in a Zambian paediatric population. Bulletin of the World Health Organization 75 | 1994 | Zambia | 894 | Africa | CS | Outp. | No | <5y | 0–5 | Unknown | 187 | 260 | 72% |
| Huebner (2000) | Prevalence of nasopharyngeal antibiotic-resistant pneumococcal carriage in children attending private paediatric practices in Johannesburg. South African Medical Journal 90 | 1999 | South Africa | 710 | Africa | CS | Outp. | No | <5y | 1–5 | Unknown | 121 | 303 | 40% |
| Hussain (2005) | A longitudinal household study of Streptococcus pneumoniae nasopharyngeal carriage in a UK setting. Epidemiology and Infection 133 (5) | 2001 | United Kingdom | 826 | Europe | CS | Unkn. | No | <5y | 0–4 | Unknown | | | Unkn. |
| | | 2001 | United Kingdom | 826 | Europe | CS | Unkn. | No | 5–17y | 5–19 | Unknown | | | Unkn. |
| | | 2001 | United Kingdom | 826 | Europe | CS | Unkn. | No | 18+y | ≥20 | Unknown | | | Unkn. |
| Ishihara (2015) | Detection and Serotyping of Streptococcus pneumoniae Carried in Healthy Adults with a Modified PCR Method. (Japanese). Kansenshogaku zasshi. The Journal of the Japanese Association for Infectious Diseases 89 (3) | 2011 | Japan | 392 | Asia | Unkn. | Unkn. | No | 18+y | ≥40 | Unknown | 36 | 110 | 33% |
| Joloba (2001) | High prevalence of carriage of antibiotic-resistant Streptococcus pneumoniae in children in Kampala Uganda. International Journal of Antimicrobial Agents 17 | 1995 | Uganda | 800 | Africa | CS | Outp. | No | <5y | 0–3 | Unknown | 118 | 191 | 62% |
| Jourdain (2011) | Differences in nasopharyngeal bacterial carriage in preschool children from different socio-economic origins. Clinical Microbiology and Infection 17 | 2006 | Belgium | 56 | Europe | CS | Unkn. | No | <5y | 3–6 | Unknown | 346 | 1347 | 26% |
| Kamng'ona (2015) | High multiple carriage and emergence of Streptococcus pneumoniae vaccine serotype variants in Malawian children. BMC Infectious Diseases 15 | 2008 | Malawi | 454 | Africa | CS | Rand. | No | <5y | 0–13 | 4.9 | 72 | | Unkn. |
| Kandasamy (2015) | Multi-serotype pneumococcal nasopharyngeal carriage prevalence in vaccine naive Nepalese children, assessed using molecular serotyping. PLOS ONE 10 (2) | 2012 | Nepal | 524 | Asia | CS | Unkn. | No | <5y | 0–2 | Unknown | 309 | 600 | 52% |
| Katsarolis (2009) | Risk factors for nasopharyngeal carriage of drug-resistant Streptococcus pneumoniae: data from a nation-wide surveillance study in Greece. BMC Infectious Diseases 9 | 2004 | Greece | 300 | Europe | CS | Unkn. | No | <5y | 0–6 | 4 | 317 | | Unkn. |
| | | 2006 | Ethiopia | 231 | Africa | CS | Trial | No | <5y | 0–9 | Unknown | 76 | 110 | 69% |

| | | | | | | | | | | | | | | |
|---|---|---|---|---|---|---|---|---|---|---|---|---|---|---|
| Keenan (2016) | Nasopharyngeal Pneumococcal Serotypes Before and After Mass Azithromycin Distributions for Trachoma. Journal of the Pediatric Infectious Diseases Society 5 (2) | 2006 | Ethiopia | 231 | Africa | CS | Trial | No | <5y | 0–9 | Unknown | 98 | 120 | 82% |
| Kellner (1999) | Streptococcus pneumoniae carriage in children attending 59 Canadian child care centers. Archives of Pediatrics and Adolescent Medicine 153 | 1995 | Canada | 124 | Americas | CS | Outp. | No | <5y | 1–2 | Unknown | 568 | 1322 | 43% |
| Kim (2011) | Nasopharyngeal Pneumococcal Carriage of Children Attending Day Care Centers in Korea: Comparison between Children Immunized with 7-valent Pneumococcal Conjugate Vaccine and Non-immunized. Journal of Korean Medical Science 26 (2) | 2008 | South Korea | 410 | Asia | CS | Unkn. | No | <5y | 1–4 | Unknown | 63 | 200 | 32% |
| Klugman (1998) | Antibiotic resistance of nasopharyngeal isolates of Streptococcus pneumoniae from children in Lesotho. Bulletin of the World Health Organization 76 | 1995 | Lesotho | 426 | Africa | CS | Rand. | No | <5y | 0–4 | Unknown | 310 | | Unkn. |
| Kobayashi (2017) | Pneumococcal carriage and antibiotic susceptibility patterns from two cross-sectional colonization surveys among children aged <5 years prior to the introduction of 10-valent pneumococcal conjugate vaccine - Kenya, 2009-2010. BMC Infectious Diseases 17 | 2009 | Kenya | 404 | Africa | CS | Rand. | No | <5y | 0–4 | Unknown | 983 | 1087 | 90% |
| Kozlov (2006) | Antimicrobial resistance of nasopharyngeal pneumococci from children from day-care centres and orphanages in Russia: results of a unique prospective multicentre study. Clinical Microbiology and Infection 12 | 2001 | Russia | 643 | Europe | CS | Unkn. | No | <5y | 0–4 | Unknown | 2056 | 4153 | 50% |
| Kumar (2014) | Nasopharyngeal carriage, antibiogram and serotype distribution of Streptococcus pneumoniae among healthy under five children. Indian Journal of Medical Research 140 (2) | 2010 | India | 356 | Asia | CS | Outp. | No | <5y | ≤5 | Unknown | 53 | 190 | 28% |
| Lagos (2008) | Age- and serotype-specific pediatric invasive pneumococcal disease: insights from systematic surveillance in Santiago, Chile, 1994–2007. Journal of Infectious Diseases 198 | 2000 | Chile | 152 | Americas | Long. | Rand. | No | <5y | 1–1 | 1 | 184 | 457 | 40% |
| | | 2001 | Chile | 152 | Americas | Long. | Rand. | No | <5y | <1 | 0 | 177 | 464 | 38% |
| | | 2001 | Chile | 152 | Americas | Long. | Rand. | No | <5y | <1 | 0 | 127 | 420 | 30% |
| | | 2001 | Chile | 152 | Americas | Long. | Rand. | No | <5y | 1–1 | 1 | 167 | 440 | 38% |
| | | 2001 | Chile | 152 | Americas | Long. | Rand. | No | <5y | 2–2 | 2 | 161 | 434 | 37% |
| Lauderdale (2005) | High carriage rate of high-level penicillin-resistant Streptococcus pneumoniae in a Taiwan kindergarten associated with a case of pneumococcal meningitis. BMC Infectious Diseases 5 (1) | 2002 | Taiwan | 158 | Asia | CS | Unkn. | No | 5–17y | 4–6 | Unknown | 31 | 77 | 40% |
| Laval (2006) | Serotypes of carriage and invasive isolates of Streptococcus pneumoniae in Brazilian children in the era of pneumococcal vaccines. Clinical Microbiology and Infection 12 (1) | 2000 | Brazil | 76 | Americas | CS | Outp. | No | <5y | <1 | 0 | 50 | 120 | 42% |
| | | 2000 | Brazil | 76 | Americas | CS | Outp. | No | <5y | 1–1 | Unknown | 75 | 182 | 41% |
| Leach (1997) | A prospective study of the impact of community-based azithromycin treatment of trachoma on carriage and resistance of Streptococcus pneumoniae. Clinical Infectious Diseases 24 | 1995 | Australia | 36 | Oceania | CS | Unkn. | Yes | 5–17y | 5–14 | Unknown | 54 | 79 | 68% |
| | | 1995 | Australia | 36 | Oceania | CS | Unkn. | Yes | 5–17y | 6–14 | Unknown | 34 | 39 | 87% |
| Leibovitz (1999) | Nasopharyngeal carriage of multidrug-resistant Streptococcus pneumoniae in institutionalized HIV-infected and HIV-negative children in northeastern Romania. International Journal of Infectious Diseases 3 (4) | 1996 | Romania | 642 | Europe | CS | Unkn. | No | <5y | 0–5 | 1 | 93 | 202 | 46% |
| Leibovitz (2007) | Antibiotic susceptibility, serotype distribution and vaccine coverage of nasopharyngeal and oropharyngeal streptococcus pneumoniae in a day-care in St. Petersburg, Russia. Scandinavian Journal of Infectious Diseases 39 | 2003 | Russia | 643 | Europe | CS | Unkn. | No | <5y | 1–5 | Unknown | 75 | 125 | 60% |
| Leino (2001) | Pneumococcal carriage in children during their first two years: important role of family exposure. Pediatric Infectious Disease Journal 20 | 1994 | Finland | 246 | Europe | Long. | Unkn. | No | <5y | 0–4 | Unknown | 189 | 676 | 28% |
| Leino (2008) | Clustering of serotypes in a longitudinal study of Streptococcus pneumoniae carriage in three day care centres. BMC Infectious Diseases 8 | 2001 | Finland | 246 | Europe | Long. | Unkn. | No | <5y | 1–6 | Unknown | 132 | 521 | 25% |
| | | 2001 | Finland | 246 | Europe | Long. | Unkn. | No | 5–17y | 5–10 | Unknown | 36 | 287 | 13% |
| | | 2001 | Finland | 246 | Europe | Long. | Unkn. | No | 18+y | ≥18 | Unknown | 35 | 1133 | 3% |
| Levidiotou (2006) | Serotype distribution of Streptococcus pneumoniae in north-western Greece and implications for a vaccination programme. FEMS Immunology and Medical Microbiology 48 (2) | 2000 | Greece | 300 | Europe | CS | Unkn. | No | <5y | 3–4 | Unknown | 172 | 464 | 37% |
| Levine (2012) | Dynamics of pneumococcal acquisition and carriage in young adults during training in confined settings in Israel. PLOS ONE 7 | 2007 | Israel | 376 | Asia | Long. | Unkn. | No | 18+y | ≥18 | Unknown | 202 | 1872 | 11% |
| Li (2001) | Nasal carriage of Streptococcus pneumoniae among children in Beijing. Chinese Medical Journal 114 (11) | 1999 | China | 156 | Asia | CS | Outp. | No | <5y | 0–5 | Unknown | 100 | 269 | 37% |
| Lin (2009) | National survey of invasive pneumococcal diseases in Taiwan under partial PCV7 vaccination in 2007: emergence of serotype 19A with high invasive potential. Vaccine 27 | 2007 | Taiwan | 158 | Asia | CS | Outp. | No | <5y | 0–5 | Unknown | 165 | 1128 | 15% |

| | | | | | | | | | | | | | | |
|---|---|---|---|---|---|---|---|---|---|---|---|---|---|---|
| Lindstrand (2016) | Pneumococcal Carriage in Children under Five Years in Uganda-Will Present Pneumococcal Conjugate Vaccines Be Appropriate?. PLOS ONE 11 (11) | 2008 | Uganda | 800 | Africa | CS | Rand. | No | <5y | 0–5 | Unknown | 957 | 1723 | 101% |
| Lloyd-Evans (1996) | Nasopharyngeal carriage of pneumococci in Gambian children and in their families. Pediatric Infectious Disease Journal 15 (10) | 1990 | Gambia | 270 | Africa | CS | Rand. | No | <5y | 0–4 | Unknown | 86 | 113 | 76% |
| Lopez (1999) | Epidemiological study of Streptococcus pneumoniae carriers in healthy primary-school children. European Journal of Clinical Microbiology and Infectious Diseases 18 (11) | 1997 | Spain | 724 | Europe | Long. | Unkn. | No | 5–17y | 6–6 | 6 | 120 | 996 | 12% |
| Lucarevschi (2003) | Oropharyngeal carriage of Streptococcus pneumoniae by children attending day care centers in Taubate, SP: Correlation between serotypes and the conjugated heptavalent pneumococcal vaccine. PortugueseColonizacao de orofaringe por Streptococcus pneumoniae em criancas de creches municipais de Taubate-SP: Correlacao entre os principais sorotipos e a vacina pneumococica conjugada heptavalente. Jornal de Pediatria 79 | 1998 | Brazil | 76 | Americas | CS | Unkn. | No | <5y | 1–6 | 5 | 209 | 987 | 21% |
| Luminos (2014) | Nasopharyngeal carriage of Streptococcus pneumoniae in Romanian children before the introduction of the pneumococcal conjugated vaccination into the national immunization programme: a national, multi-centre, cross-sectional observational study. International Journal of Infectious Diseases 29 | 2011 | Romania | 642 | Europe | CS | Outp. | No | <5y | 0–1 | Unknown | 31 | 186 | 17% |
| | | 2011 | Romania | 642 | Europe | CS | Outp. | No | <5y | 1–2 | Unknown | 162 | 751 | 22% |
| | | 2011 | Romania | 642 | Europe | CS | Outp. | No | <5y | 3–4 | Unknown | 312 | 1063 | 29% |
| Madhi (2007) | Long-term effect of pneumococcal conjugate vaccine on nasopharyngeal colonization by Streptococcus pneumoniae–and associated interactions with Staphylococcus aureus and Haemophilus influenzae colonization–in HIV-Infected and HIV-uninfected children. Journal of Infectious Diseases 196 (11) | 2001 | South Africa | 710 | Africa | CS | Trial | No | 5–17y | 5–6 | 5 | 79 | 150 | 53% |
| Madhi (2013) | Acquisition of Streptococcus pneumoniae in pneumococcal conjugate vaccine-naive South African children and their mothers. Pediatric Infectious Disease Journal 32 | 2007 | South Africa | 710 | Africa | Long. | Outp. | No | <5y | 1–1 | 0 | 1093 | 1851 | 59% |
| | | 2007 | South Africa | 710 | Africa | Long. | Outp. | No | 18+y | ≥18 | 28 | 343 | 1846 | 19% |
| Mahanta (2012) | Serotype distribution and sensitivity pattern of nasopharyngeal colonizing streptococcus pneumoniae among rural children of Eastern India. Indian Journal of Medical Research 136 | 2009 | India | 356 | Asia | CS | Unkn. | No | <5y | 0–4 | Unknown | 104 | 811 | 13% |
| Malfroot (2004) | A cross-sectional survey of the prevalence of Streptococcus pneumoniae nasopharyngeal carriage in Belgian infants attending day care centres. Clinical Microbiology and Infection 10 | 2000 | Belgium | 56 | Europe | CS | Unkn. | No | <5y | 1–3 | Unknown | 98 | 467 | 21% |
| Masri (2013) | Determination of phenotypes and pneumococcal surface protein a family types of Streptococcus pneumoniae from Malaysian healthy children. Journal of Microbiology, Immunology and Infection 46 | 2010 | Malaysia | 458 | Asia | CS | Unkn. | No | <5y | 0–5 | Unknown | 69 | 195 | 35% |
| Mastro (1993) | Use of nasopharyngeal isolates of Streptococcus pneumoniae and Haemophilus influenzae from children in Pakistan for surveillance for antimicrobial resistance. Pediatric Infectious Disease Journal 12 (10) | 1989 | Pakistan | 586 | Asia | CS | Rand. | No | <5y | 0–4 | 1 | 244 | 418 | 58% |
| Mbelle (1999) | Immunogenicity and impact on nasopharyngeal carriage of a nonvalent pneumococcal conjugate vaccine. Journal of Infectious Diseases 180 (4) | NA | South Africa | 710 | Africa | CS | Trial | No | <5y | <1 | Unknown | | 239 | Unkn. |
| Mbelle (1999) | Streptococcus pneumoniae and haemophilus influenzae type B carriage in infants presenting to Zola Community Health centre for routine immunization. Doctoral dissertation, University of the Witwatersrand | 1996 | South Africa | 710 | Africa | CS | Outp. | No | <5y | 0–5 | Unknown | 121 | 278 | 44% |
| McGee (2001) | Prevalence of serotypes and molecular epidemiology of Streptococcus pneumoniae strains isolated from children in Beijing, China: identification of two novel multiply-resistant clones. Microbial Drug Resistance 7 | 1997 | China | 156 | Asia | CS | Outp. | No | <5y | 0–5 | Unknown | 326 | | Unkn. |
| Menezes (2016) | Nasopharyngeal carriage of Streptococcus pneumoniae among children in an urban setting in Brazil prior to PCV10 introduction. Vaccine 34 (6) | 2008 | Brazil | 76 | Americas | Long. | Rand. | No | <5y | 0–5 | Unknown | 398 | 721 | 55% |
| Mercado (2012) | Pneumococcal serotypes in carrier children prior to the introduction of anti-pneumococcal vaccines in Peru (Spanish). Revista Peruana de Medicina Experimental y Salud Publica 29 (1) | 2003 | Peru | 604 | Americas | CS | Outp. | No | <5y | 0–1 | 1 | 80 | 270 | 30% |
| | | 2007 | Peru | 604 | Americas | CS | Outp. | No | <5y | <1 | Unknown | 286 | 1145 | 25% |
| Millar (2008) | Indirect effect of 7-valent pneumococcal conjugate vaccine on pneumococcal colonization among unvaccinated household members. Clinical Infectious Diseases 47 | 2001 | United States | 840 | Americas | Long. | Trial | Yes | <5y | 0–1 | 1 | 19 | 37 | 51% |
| | | 2001 | United States | 840 | Americas | Long. | Trial | Yes | <5y | 2–4 | 3 | 40 | 62 | 65% |
| | | 2001 | United States | 840 | Americas | Long. | Trial | Yes | 5–17y | 5–17 | Unknown | 210 | 568 | 37% |
| | | 2001 | United States | 840 | Americas | Long. | Trial | Yes | 18+y | 18–64 | Unknown | 54 | 677 | 8% |
| | | 2001 | United States | 840 | Americas | Long. | Trial | Yes | 18+y | 65–88 | Unknown | 5 | 29 | 17% |

| | | | | | | | | | | | | | | |
|---|---|---|---|---|---|---|---|---|---|---|---|---|---|---|
| Mills (2015) | Epidemiology of pneumococcal carriage in children under five years of age in Accra, Ghana. Infectious Diseases 47 (5) | 2011 | Ghana | 288 | Africa | CS | Rand. | No | <5y | 0.25–5 | Unknown | 207 | 423 | 49% |
| Miranda Novales (1997) | Streptococcus pneumoniae: low frequency of penicillin resistance and high resistance to trimethoprim-sulfamethoxazole in nasopharyngeal isolates from children in a rural area in Mexico. Archives of Medical Research 28 (4) | 1994 | Mexico | 484 | Americas | CS | Outp. | No | <5y | 0–4 | Unknown | 65 | 127 | 51% |
| Moll (2008) | Factors associated with pneumococcal carriage in healthy Dutch infants: the generation R study. Journal of Pediatrics 153 | 2003 | Netherlands | 528 | Europe | Long. | Rand. | No | <5y | <1 | 0 | 52 | 627 | 8% |
| | | 2003 | Netherlands | 528 | Europe | Long. | Rand. | No | <5y | <1 | 0 | 260 | 832 | 31% |
| | | 2003 | Netherlands | 528 | Europe | Long. | Rand. | No | <5y | 1–1 | 1 | 337 | 757 | 45% |
| Moniri (2014) | Serotyping, Antibiotic Susceptibility and Related Risk Factors Aspects of Nasopharyngeal Carriage of Streptococcus pneumoniae in Healthy School Students. Iranian Journal of Public Health 43 | 2011 | Iran | 364 | Asia | CS | Unkn. | No | 5–17y | 7–19 | Unknown | 291 | 2100 | 14% |
| Moreno (2013) | Changes in Streptococcus pneumoniae serotype distribution in invasive disease and nasopharyngeal carriage after the heptavalent pneumococcal conjugate vaccine introduction in Bogota, Colombia. Vaccine 31 | 2005 | Colombia | 170 | Americas | CS | Rand. | No | <5y | 1–1 | 1 | 137 | 246 | 56% |
| Mousavi (2013) | Serotyping of Streptococcus pneumoniae isolated from Tehran by Multiplex PCR: Are serotypes of clinical and carrier isolates identical?. Iranian Journal of Microbiology 5 | 2011 | Iran | 364 | Asia | CS | Outp. | No | <5y | 0–4 | Unknown | 40 | 150 | 27% |
| Moyo (2012) | Penicillin resistance and serotype distribution of Streptococcus pneumoniae in nasopharyngeal carrier children under 5 years of age in Dar es Salaam, Tanzania. Journal of Medical Microbiology 61 | 2010 | Tanzania | 834 | Africa | CS | Outp. | No | <5y | <1 | Unknown | 27 | 63 | 43% |
| | | 2010 | Tanzania | 834 | Africa | CS | Outp. | No | <5y | 1–1 | 1 | 28 | 73 | 38% |
| | | 2010 | Tanzania | 834 | Africa | CS | Outp. | No | <5y | 2–4 | Unknown | 50 | 164 | 30% |
| Müller-Graf (1999) | Population biology of Streptococcus pneumoniae isolated from oropharyngeal carriage and invasive disease. Microbiology 145 (11) | 2006 | Poland | 616 | Europe | CS | Unkn. | No | <5y | 2–5 | Unknown | 13 | 95 | 14% |
| Nackers (2017) | Carriage prevalence and serotype distribution of Streptococcus pneumoniae prior to 10-valent pneumococcal vaccine introduction: A population-based cross-sectional study in South Western Uganda, 2014. Vaccine 35 (39) | 2014 | Uganda | 800 | Africa | CS | Rand. | No | <5y | 0–1 | Unknown | 86 | 205 | 42% |
| | | 2014 | Uganda | 800 | Africa | CS | Rand. | No | <5y | 1–2 | Unknown | 78 | 182 | 43% |
| | | 2014 | Uganda | 800 | Africa | CS | Rand. | No | <5y | 2–4 | Unknown | 74 | 217 | 34% |
| | | 2014 | Uganda | 800 | Africa | CS | Rand. | No | 5–17y | 5–14 | Unknown | 68 | 417 | 16% |
| | | 2014 | Uganda | 800 | Africa | CS | Rand. | No | 18+y | ≥30 | Unknown | 2 | 208 | 1% |
| | | 2014 | Uganda | 800 | Africa | CS | Rand. | No | 18+y | 15–29 | Unknown | 1 | 117 | 1% |
| Neves (2013) | Nasopharyngeal carriage, serotype distribution and antimicrobial resistance of Streptococcus pneumoniae among children from Brazil before the introduction of the 10-valent conjugate vaccine. BMC Infectious Diseases 13 | 2010 | Brazil | 76 | Americas | CS | Outp. | No | <5y | <1 | 0 | 11 | 37 | 30% |
| | | 2010 | Brazil | 76 | Americas | CS | Outp. | No | <5y | 1–1 | 1 | 20 | 38 | 53% |
| | | 2010 | Brazil | 76 | Americas | CS | Outp. | No | <5y | 2–4 | Unknown | 74 | 124 | 60% |
| | | 2010 | Brazil | 76 | Americas | CS | Outp. | No | <5y | 5–5 | 5 | 16 | 43 | 37% |
| Neves (2018) | Population structure of Streptococcus pneumoniae colonizing children before and after universal use of pneumococcal conjugate vaccines in Brazil: Emergence and expansion of the MDR serotype 6C-CC386 lineage. Journal of Antimicrobial Chemotherapy 73 (5) | 2010 | Brazil | 76 | Americas | CS | Unkn. | No | <5y | 0–6 | 3 | | 102 | Unkn. |
| Niedzielski (2012) | Distribution of vaccine serotypes among Streptococcus pneumoniae colonizing the upper respiratory tract in healthy pre-school children in south-east Poland. Otolaryngologia Polska 66 (6) | 2002 | Poland | 616 | Europe | Long. | Unkn. | No | <5y | 3–5 | Unknown | 342 | 933 | 37% |
| Nissinen (1995) | Antimicrobial resistance of Streptococcus pneumoniae in Finland, 1987-1990. Clinical Infectious Diseases 20 (5) | 1987 | Finland | 246 | Europe | CS | Outp. | No | <5y | 3–6 | Unknown | | | Unkn. |
| O'Brien (2006) | Effect of community-wide conjugate pneumococcal vaccine use in infancy on nasopharyngeal carriage through 3 years of age: a cross-sectional study in a high-risk population. Clinical Infectious Diseases 43 | 1997 | United States | 840 | Americas | Long. | Trial | Yes | <5y | 0–1 | 1 | 6 | 8 | 75% |
| | | 1997 | United States | 840 | Americas | Long. | Trial | Yes | <5y | 2–4 | Unknown | 163 | 262 | 62% |
| | | 1998 | United States | 840 | Americas | Long. | Trial | Yes | <5y | <1 | 0 | 175 | 262 | 67% |
| | | 1998 | United States | 840 | Americas | Long. | Trial | Yes | <5y | 0–6 | Unknown | 63 | 96 | 66% |
| | | 1998 | United States | 840 | Americas | Long. | Trial | Yes | <5y | 1–1 | 1 | 147 | 239 | 62% |
| | | 1998 | United States | 840 | Americas | Long. | Trial | Yes | <5y | 1–1 | 1 | 154 | 250 | 62% |
| | | 1998 | United States | 840 | Americas | Long. | Trial | Yes | <5y | 1–6 | Unknown | 79 | 120 | 66% |

| | | | | | | | | | | | | | | |
|---|---|---|---|---|---|---|---|---|---|---|---|---|---|---|
| O’Brien (2007) | Effect of pneumococcal conjugate vaccine on nasopharyngeal colonization among immunized and unimmunized children in a community-randomized trial. Journal of Infectious Diseases 196 | | | | | | | | | | | | | |
| | | 1998 | United States | 840 | Americas | Long. | Trial | Yes | <5y | 1–6 | Unknown | 58 | 100 | 58% |
| | | 1999 | United States | 840 | Americas | Long. | Trial | Yes | <5y | <1 | Unknown | 113 | 280 | 40% |
| Ochoa (2005) | Penicillin resistance and serotypes/serogroups of Streptococcus pneumoniae in nasopharyngeal carrier children younger than 2 years in Lima, Peru. Diagnostic Microbiology and Infectious Disease 52 | 1997 | Peru | 604 | Americas | CS | Outp. | No | <5y | <1 | Unknown | 66 | 170 | 39% |
| | | 2001 | Peru | 604 | Americas | CS | Outp. | No | <5y | <1 | 0 | 65 | 226 | 29% |
| | | 2003 | Peru | 604 | Americas | CS | Outp. | No | <5y | 0–1 | 1 | 80 | 270 | 30% |
| Otsuka (2013) | Individual Risk Factors Associated With Nasopharyngeal Colonization With Streptococcus pneumoniae and Haemophilus influenzae: A Japanese Birth Cohort Study. Pediatric Infectious Disease Journal 32 | 2008 | Japan | 392 | Asia | Long. | Outp. | No | <5y | <1 | 0 | 121 | 334 | 36% |
| | | 2008 | Japan | 392 | Asia | Long. | Outp. | No | <5y | <1 | 0 | 92 | 334 | 28% |
| | | 2008 | Japan | 392 | Asia | Long. | Outp. | No | <5y | <1 | 0 | 59 | 341 | 17% |
| | | 2008 | Japan | 392 | Asia | Long. | Outp. | No | <5y | 1–1 | 1.5 | 156 | 324 | 48% |
| | | 2008 | Japan | 392 | Asia | Long. | Outp. | No | <5y | 3–3 | 3 | 86 | 191 | 45% |
| Ousmane (2017) | Serotype Distribution and Antimicrobial Sensitivity Profile of Streptococcus pneumoniae Carried in Healthy Toddlers before PCV13 Introduction in Niamey, Niger. PLOS ONE 12 (1) | 2007 | Niger | 562 | Africa | CS | Outp. | No | <5y | 0–2 | Unknown | 654 | 1200 | 55% |
| Palmu (2012) | A longitudinal study of Streptococcus pneumoniae carriage in a cohort of infants and their mothers on the Thailand-Myanmar border. Scandinavian Journal of Infectious Diseases 44 (6) | 2007 | Thailand | 764 | Asia | Long. | Outp. | No | <5y | <1 | 0 | 91 | 232 | 39% |
| Palmu (2012) | Nasopharyngeal carriage of Streptococcus pneumoniae and pneumococcal urine antigen test in healthy elderly subjects. Scandinavian Journal of Infectious Diseases 44 (6) | 2003 | Finland | 246 | Europe | CS | Unkn. | No | 18+y | 65–92 | 70 | 31 | 590 | 5% |
| Paraskakis (2006) | Serotypes and antimicrobial susceptibilities of 1033 pneumococci isolated from children in Greece during 2001-2004. Clinical Microbiology and Infection 12 (5) | 2003 | Greece | 300 | Europe | CS | Unkn. | No | <5y | 3–6 | Unknown | 206 | 873 | 24% |
| Park (2008) | Impact of conjugate vaccine on transmission of antimicrobial-resistant Streptococcus pneumoniae among Alaskan children. Pediatric Infectious Disease Journal 27 | 2000 | United States | 840 | Americas | CS | Outp. | No | <5y | <1 | Unknown | 37 | 108 | 34% |
| | | 2000 | United States | 840 | Americas | CS | Outp. | No | <5y | 1–1 | Unknown | 56 | 127 | 44% |
| | | 2000 | United States | 840 | Americas | CS | Outp. | No | <5y | 2–4 | Unknown | 79 | 216 | 37% |
| Peñuela (1999) | Colonizacion nasofarigea y resistencia antimicrobiana de Streptococcus pneumoniae en ninos de una guarderia en Santa Fe de Bogota. Biomedica 19 | 1997 | Colombia | 170 | Americas | Long. | Unkn. | No | <5y | 0–1 | Unknown | 51 | 82 | 62% |
| Petrosillo (2002) | Prevalence, determinants, and molecular epidemiology of Streptococcus pneumoniae isolates colonizing the nasopharynx of healthy children in Rome. European Journal of Clinical Microbiology and Infectious Diseases 21 | 1999 | Italy | 380 | Europe | CS | Unkn. | No | <5y | 0–5 | Unknown | 91 | 610 | 15% |
| Principi (2002) | Nasopharyngeal carriage of Streptococcus pneumoniae in healthy children: implications for the use of heptavalent pneumococcal conjugate vaccine. Emerging Infectious Diseases 8 | 2000 | Italy | 380 | Europe | CS | Unkn. | No | <5y | 0–6 | Unknown | 242 | 2799 | 9% |
| Prymula (2009) | Effect of vaccination with pneumococcal capsular polysaccharides conjugated to Haemophilus influenzae-derived protein D on nasopharyngeal carriage of Streptococcus pneumoniae and H. influenzae in children under 2 years of age. Vaccine 28 | 2004 | Czechia | 203 | Europe | CS | Trial | No | <5y | 0–2 | 1 | 39 | 175 | 22% |
| Prymula (2011) | Impact of the 10-valent pneumococcal non-typeable Haemophilus influenzae Protein D conjugate vaccine (PHiD-CV) on bacterial nasopharyngeal carriage. Vaccine 29 | 2007 | Czechia | 203 | Europe | CS | Trial | No | <5y | 1–1 | 1 | 161 | 336 | 48% |
| Raman (2017) | Demographic profile of healthy children with nasopharyngeal colonisation of Streptococcus pneumoniae: A research paper. Indian Journal of Medical Microbiology 35 (4) | 2012 | India | 356 | Asia | CS | Outp. | No | <5y | 0.5–5 | Unknown | 45 | 450 | 10% |
| Rashid (2013) | Oro-pharyngeal carriage and antimicrobial susceptibility of streptococcus pneumoniae from healthy children. Internet Journal of Microbiology 11 (1) | 2010 | Malaysia | 458 | Asia | CS | Unkn. | No | <5y | 1–2 | Unknown | 2 | 65 | 3% |
| Raymond (2000) | Sequential colonization by Streptococcus pneumoniae of healthy children living in an orphanage. Journal of Infectious Diseases 181 (6) | 1996 | France | 250 | Europe | Long. | Unkn. | No | <5y | 0–1 | Unknown | 167 | 289 | 58% |
| Raymond (2002) | Factors influencing Streptococcus pneumoniae carriage. Medecine et Maladies Infectieuses 32 | 1996 | France | 250 | Europe | Long. | Unkn. | No | <5y | 0–2 | Unknown | 111 | 852 | 13% |
| Regev-Yochay (2004) | Nasopharyngeal carriage of Streptococcus pneumoniae by adults and children in community and family settings. Clinical Infectious Diseases 38 | 2001 | Israel | 376 | Asia | CS | Outp. | No | <5y | 0–6 | Unknown | 214 | 404 | 53% |
| | | 2001 | Israel | 376 | Asia | CS | Outp. | No | 18+y | ≥18 | Unknown | 48 | 1300 | 4% |
| | | 2009 | Palestinian Territories | 275 | Asia | CS | Rand. | Yes | <5y | 0–5 | 2 | 189 | 379 | 50% |

| | | | | | | | | | | | | | | |
|---|---|---|---|---|---|---|---|---|---|---|---|---|---|---|
| Regev-Yochay (2012) | Streptococcus pneumoniae carriage in the Gaza strip. PLOS ONE 7 | 2009 | Palestinian Territories | 275 | Asia | CS | Rand. | Yes | 18+y | ≥18 | Unknown | 34 | 379 | 9% |
| Reis (2008) | Transmission of Streptococcus pneumoniae in an urban slum community. Journal of Infection 57 | 2000 | Brazil | 76 | Americas | CS | Unkn. | No | <5y | <1 | Unknown | 6 | 8 | 75% |
| | | 2000 | Brazil | 76 | Americas | CS | Unkn. | No | <5y | 1–1 | Unknown | 4 | 7 | 57% |
| | | 2000 | Brazil | 76 | Americas | CS | Unkn. | No | <5y | 2–4 | Unknown | 23 | 35 | 66% |
| | | 2000 | Brazil | 76 | Americas | CS | Unkn. | No | <5y | Unreported | Unknown | 19 | 117 | 16% |
| | | 2000 | Brazil | 76 | Americas | CS | Unkn. | No | 5–17y | 5–17 | Unknown | 52 | 95 | 55% |
| Rey (2002) | Antimicrobial susceptibility and serotypes of nasopharyngeal Streptococcus pneumoniae in children with pneumonia and in children attending day-care centres in Fortaleza, Brazil. International Journal of Antimicrobial Agents 20 | 1998 | Brazil | 76 | Americas | CS | Unkn. | No | <5y | 1–4 | Unknown | 269 | 374 | 72% |
| Roca (2006) | Serotype distribution and antibiotic susceptibility of invasive and nasopharyngeal isolates of Streptococcus pneumoniae among children in rural Mozambique. Tropical Medicine and International Health 11 | 2003 | Mozambique | 508 | Africa | CS | Outp. | No | <5y | 0–4 | 1 | 192 | 248 | 77% |
| Roca (2011) | Effects of community-wide vaccination with PCV-7 on pneumococcal nasopharyngeal carriage in the Gambia: a cluster-randomized trial. PLOS Medicine 8 | 2003 | Gambia | 270 | Africa | CS | Trial | No | <5y | 2–4 | 3 | 197 | 219 | 90% |
| | | 2003 | Gambia | 270 | Africa | CS | Trial | No | 5–17y | 5–17 | Unknown | 741 | 896 | 83% |
| | | 2003 | Gambia | 270 | Africa | CS | Trial | No | 18+y | ≥60 | Unknown | 106 | 206 | 51% |
| | | 2003 | Gambia | 270 | Africa | CS | Trial | No | 18+y | 18–59 | Unknown | 444 | 773 | 57% |
| Roche (2007) | Prevalence of nasopharyngeal carriage of pneumococcus in preschool children attending day care in London. Archives of Disease in Childhood 92 | 2003 | United Kingdom | 826 | Europe | CS | Unkn. | No | <5y | <1 | Unknown | | | Unkn. |
| | | 2003 | United Kingdom | 826 | Europe | CS | Unkn. | No | <5y | 1–1 | 1 | | | Unkn. |
| | | 2003 | United Kingdom | 826 | Europe | CS | Unkn. | No | <5y | 2–4 | Unknown | | | Unkn. |
| Ronchetti (1999) | Resistance patterns of Streptococcus pneumoniae from children in central Italy. European Journal of Clinical Microbiology and Infectious Diseases 18 (5) | 1997 | Italy | 380 | Europe | CS | Outp. | No | <5y | 3–5 | Unknown | 121 | 1146 | 11% |
| Rosen (1996) | Antibodies to pneumococcal polysaccharides in human milk: lack of relationship to colonization and acute otitis media. Pediatric Infectious Disease Journal 15 (6) | 1995 | Sweden | 752 | Europe | Long. | Rand. | No | <5y | <1 | Unknown | 113 | 423 | 27% |
| | | 1995 | Sweden | 752 | Europe | Long. | Rand. | No | <5y | <1 | 0 | 52 | 448 | 12% |
| | | 1995 | Sweden | 752 | Europe | Long. | Rand. | No | <5y | <1 | Unknown | 115 | 404 | 28% |
| Rowe (2000) | Antimicrobial resistance of nasopharyngeal isolates of Streptococcus pneumoniae and Haemophilus influenzae from children in the Central African Republic. Pediatric Infectious Disease Journal 19 | 1995 | Central African Republic | 140 | Africa | CS | Outp. | No | <5y | <1 | 0 | 32 | 39 | 82% |
| | | 1995 | Central African Republic | 140 | Africa | CS | Outp. | No | <5y | 1–1 | Unknown | 27 | 36 | 75% |
| | | 1995 | Central African Republic | 140 | Africa | CS | Outp. | No | <5y | 2–4 | Unknown | 23 | 31 | 74% |
| Rusen (1997) | Nasopharyngeal pneumococcal colonization among Kenyan children: antibiotic resistance, strain types and associations with human immunodeficiency virus type 1 infection. Pediatric Infectious Disease Journal 16 (7) | 1990 | Kenya | 404 | Africa | CS | Outp. | No | <5y | 0–5 | Unknown | 92 | 207 | 44% |
| Russell (2006) | Pneumococcal nasopharyngeal carriage and patterns of penicillin resistance in young children in Fiji. Annals of Tropical Paediatrics 26 (3) | 2003 | Fiji | 242 | Oceania | CS | Outp. | No | <5y | 0–1 | 0 | 195 | 440 | 44% |
| Russell (2010) | Pneumococcal nasopharyngeal carriage following reduced doses of a 7-valent pneumococcal conjugate vaccine and a 23-valent pneumococcal polysaccharide vaccine booster. Clinical and Vaccine Immunology 17 | 2004 | Fiji | 242 | Oceania | Long. | Trial | No | <5y | <1 | 0 | 54 | 120 | 45% |
| | | 2004 | Fiji | 242 | Oceania | Long. | Trial | No | <5y | <1 | Unknown | 49 | 121 | 40% |
| | | 2004 | Fiji | 242 | Oceania | Long. | Trial | No | <5y | 1–1 | 1 | 50 | 122 | 41% |
| Sá-Leao (2008) | High rates of transmission of and colonization by Streptococcus pneumoniae and Haemophilus influenzae within a day care center revealed in a longitudinal study. Journal of Clinical Microbiology 46 (1) | 1998 | Portugal | 620 | Europe | Long. | Unkn. | No | <5y | 1–3 | Unknown | 254 | 414 | 61% |
| Saeed (2011) | Carriage rates, circulating serotypes and antibiotic resistance among Streptococcus pneumoniae in healthy infants in Yei, South Sudan. South Sudan Medical Journal 4 | 2000 | South Sudan | 728 | Africa | CS | Outp. | No | <5y | 0–1 | Unknown | 14 | 38 | 37% |

| | | | | | | | | | | | | | | |
|---|---|---|---|---|---|---|---|---|---|---|---|---|---|---|
| Safari (2012) | Nasopharyngeal colonization of Streptococcus pneumoniae in elderly people, in Jakarta, Indonesia. International Journal of Infectious Diseases 16 | 2010 | Indonesia | 360 | Asia | CS | Outp. | No | 18+y | ≥60 | Unknown | 4 | 155 | 3% |
| Saha (2003) | Comparison of antibiotic resistance and serotype composition of carriage and invasive pneumococci among Bangladeshi children: implications for treatment policy and vaccine formulation. Journal of Clinical Microbiology 41 | 1999 | Bangladesh | 50 | Asia | CS | Rand. | No | <5y | 0–5 | Unknown | 1301 | 2831 | 46% |
| Salt (2007) | Social mixing with other children during infancy enhances antibody response to a pneumococcal conjugate vaccine in early childhood. Clinical and Vaccine Immunology 14 (5) | 2003 | United Kingdom | 826 | Europe | CS | Rand. | No | <5y | 1–1 | Unknown | 99 | 159 | 62% |
| Salter (2017) | A longitudinal study of the infant nasopharyngeal microbiota: The effects of age, illness and antibiotic use in a cohort of South East Asian children. PLOS Neglected Tropical Diseases 11 (10) | 2007 | Thailand | 764 | Asia | Long. | Unkn. | No | <5y | 0–2 | 1 | 468 | 517 | 91% |
| Sanders (2009) | Effect of reduced-dose schedules with 7-valent pneumococcal conjugate vaccine on nasopharyngeal pneumococcal carriage in children: A randomized controlled trial. Journal of the American Medical Association 302 | 2005 | Netherlands | 528 | Europe | Long. | Trial | No | <5y | 1–1 | 1 | 214 | 319 | 67% |
| | | 2005 | Netherlands | 528 | Europe | Long. | Trial | No | <5y | 1–1 | 1 | 215 | 317 | 68% |
| | | 2005 | Netherlands | 528 | Europe | Long. | Trial | No | <5y | 1–2 | Unknown | 211 | 321 | 66% |
| Sandgen (2004) | Effect of clonal and serotype-specific properties on the invasive capacity of Streptococcus pneumoniae. Journal of Infectious Diseases 189 (5) | 1997 | Sweden | 752 | Europe | CS | Unkn. | No | <5y | 1–6 | Unknown | 246 | 611 | 40% |
| Schaumburg (2013) | Streptococcus pneumoniae colonization in remote African Pygmies. Transactions of the Royal Society of Tropical Medicine and Hygiene 107 | 2011 | Gabon | 266 | Africa | CS | Rand. | Yes | <5y | <1 | Unknown | 0 | 1 | 0% |
| | | 2011 | Gabon | 266 | Africa | CS | Rand. | Yes | <5y | 1–1 | Unknown | 0 | 1 | 0% |
| | | 2011 | Gabon | 266 | Africa | CS | Rand. | Yes | <5y | 2–4 | Unknown | 15 | 19 | 79% |
| | | 2011 | Gabon | 266 | Africa | CS | Rand. | Yes | 5–17y | 5–17 | Unknown | 14 | 32 | 44% |
| | | 2011 | Gabon | 266 | Africa | CS | Rand. | Yes | 18+y | ≥60 | Unknown | 2 | 5 | 40% |
| | | 2011 | Gabon | 266 | Africa | CS | Rand. | Yes | 18+y | 18–59 | Unknown | 7 | 45 | 16% |
| Scott (2008) | The descriptive epidemiology of Streptococcus pneumoniae and Haemophilus influenzae nasopharyngeal carriage in children and adults in Kilifi district, Kenya. Pediatric Infectious Disease Journal 27 | 2004 | Kenya | 404 | Africa | CS | Rand. | No | <5y | 0–4 | 2 | 198 | 349 | 57% |
| Scott (2012) | Rates of acquisition of pneumococcal colonization and transmission probabilities, by serotype, among newborn infants in Kilifi District, Kenya. Clinical Infectious Diseases 55 | 2006 | Kenya | 404 | Africa | Long. | Outp. | No | 18+y | ≥18 | Unknown | 205 | 2428 | 8% |
| Scott (2012) | The prevalence and risk factors for pneumococcal colonization of the nasopharynx among children in Kilifi District, Kenya. PLOS ONE 7 | 2006 | Kenya | 404 | Africa | CS | Rand. | No | <5y | 1–4 | Unknown | 1868 | 2840 | 66% |
| Sener (1998) | Rate of carriage, serotype distribution and penicillin resistance of Streptococcus pneumoniae in healthy children. Zentralblatt fur Bakteriologie 288 (3) | 1995 | Turkey | 792 | Asia | CS | Outp. | No | <5y | 1–4 | Unknown | 71 | 248 | 29% |
| Shaumburg (2013) | Carriage of encapsulated bacteria in Gabonese children with sickle cell anaemia. Clinical Microbiology and Infection 19 | 2010 | Gabon | 266 | Africa | CS | Outp. | No | <5y | <1 | Unknown | 0 | 2 | 0% |
| | | 2010 | Gabon | 266 | Africa | CS | Outp. | No | <5y | 1–1 | Unknown | 2 | 8 | 25% |
| | | 2010 | Gabon | 266 | Africa | CS | Outp. | No | <5y | 2–4 | Unknown | 9 | 42 | 21% |
| | | 2010 | Gabon | 266 | Africa | CS | Outp. | No | 5–17y | 5–17 | Unknown | 8 | 90 | 9% |
| Simões (2011) | Clonal evolution leading to maintenance of antibiotic resistance rates among colonizing pneumococci in the PCV7 era in Portugal. Journal of Clinical Microbiology 49 (8) | 2001 | Portugal | 620 | Europe | CS | Unkn. | No | <5y | <1 | 0 | 22 | 30 | 73% |
| | | 2001 | Portugal | 620 | Europe | CS | Unkn. | No | <5y | 1–1 | 1 | 45 | 60 | 75% |
| | | 2001 | Portugal | 620 | Europe | CS | Unkn. | No | <5y | 2–4 | Unknown | 259 | 399 | 65% |
| | | 2001 | Portugal | 620 | Europe | CS | Unkn. | No | 5–17y | 5–6 | Unknown | 139 | 228 | 61% |
| Skovberg (2013) | Low rate of pneumococci non-susceptible to penicillin in healthy Swedish toddlers. Scandinavian Journal of Infectious Diseases 45 | 2004 | Sweden | 752 | Europe | CS | Outp. | No | <5y | 1–1 | Unknown | 297 | 663 | 45% |
| Sleeman (2005) | Acquisition of Streptococcus pneumoniae and nonspecific morbidity in infants and their families: a cohort study. Pediatric Infectious Disease Journal 24 (2) | 1999 | United Kingdom | 826 | Europe | Long. | Rand. | No | <5y | <1 | Unknown | 518 | 1907 | 27% |
| Slotved (2013) | Penicillin resistance and serotype distribution of Streptococcus pneumoniae in Ghanaian children less than six years of age. BMC Infectious Diseases 13 | 2011 | Ghana | 288 | Africa | CS | Unkn. | No | <5y | <1 | 0 | 6 | 7 | 86% |
| | | 2011 | Ghana | 288 | Africa | CS | Unkn. | No | <5y | 1–1 | 1 | 33 | 73 | 45% |
| | | 2011 | Ghana | 288 | Africa | CS | Unkn. | No | <5y | 2–5 | 3 | 232 | 341 | 68% |

| | | | | | | | | | | | | | | |
|---|---|---|---|---|---|---|---|---|---|---|---|---|---|---|
| Sluijter (1998) | Molecular characterization of pneumococcal nasopharynx isolates collected from children during their first 2 years of life. Journal of Clinical Microbiology 36 (8) | 1995 | Netherlands | 528 | Europe | Long. | Rand. | No | <5y | 0–2 | Unknown | 58 | 247 | 23% |
| Solorzano (2005) | Prevalence of Streptococcus pneumoniae serotypes on nasopharyngeal colonization in children of Mexico City. SpanishSerotipos prevalantes de Streptococcus pneumoniae colonizadores de nasofaringe, en ninos del Distrito Federal. Salud Publica de Mexico 47 | 2000 | Mexico | 484 | Americas | CS | Rand. | No | <5y | 0–5 | Unknown | 122 | 573 | 21% |
| Song (2001) | Carriage of antibiotic-resistant pneumococci among Asian children: a multinational surveillance by the Asian Network for Surveillance of Resistant Pathogens (ANSORP). Clinical Infectious Diseases 32 | 1998 | China | 156 | Asia | CS | Rand. | No | <5y | 0–4 | Unknown | 95 | 267 | 36% |
| | | 1998 | India | 356 | Asia | CS | Rand. | No | <5y | 0–4 | Unknown | 77 | 227 | 34% |
| | | 1998 | Malaysia | 458 | Asia | CS | Rand. | No | <5y | 0–4 | Unknown | 58 | 762 | 8% |
| | | 1998 | Philippines | 608 | Asia | CS | Rand. | No | <5y | 0–4 | Unknown | 95 | 307 | 31% |
| | | 1998 | Saudi Arabia | 682 | Asia | CS | Rand. | No | <5y | 0–4 | Unknown | 26 | 830 | 3% |
| | | 1998 | Singapore | 702 | Asia | CS | Rand. | No | <5y | 0–4 | Unknown | 41 | 491 | 8% |
| | | 1998 | South Korea | 410 | Asia | CS | Rand. | No | <5y | 0–4 | Unknown | 62 | 500 | 12% |
| | | 1998 | Sri Lanka | 144 | Asia | CS | Rand. | No | <5y | 0–4 | Unknown | 16 | 493 | 3% |
| | | 1998 | Taiwan | 158 | Asia | CS | Rand. | No | <5y | 0–4 | Unknown | 23 | 288 | 8% |
| | | 1998 | Thailand | 764 | Asia | CS | Rand. | No | <5y | 0–4 | Unknown | 165 | 503 | 33% |
| | | 1998 | Vietnam | 704 | Asia | CS | Rand. | No | <5y | 0–4 | Unknown | 92 | 295 | 31% |
| Soraa (2011) | Study of nasopharyngeal colonization by Streptococcus pneumoniae and its antibiotics resistance in healthy children less than 2 years of age in the Marrakech region (Morocco). (French). Archives de Pediatrie 18 | 2008 | Morocco | 504 | Africa | CS | Outp. | No | <5y | 0–1 | 1 | 302 | 660 | 46% |
| Stubbs (2005) | Streptococcus pneumoniae and noncapsular Haemophilus influenzae nasal carriage and hand contamination in children - A comparison of two populations at risk of otitis media. Pediatric Infectious Disease Journal 24 (5) | NA | Australia | 36 | Oceania | CS | Rand. | Yes | <5y | 0–3 | Unknown | | 294 | Unkn. |
| Sulikowska (2003) | The carriage of Streptococcus pneumoniae in the nasopharynx of children under 5 years of age in selected settings in Warsaw. PolishNosicielstwo nosogardlowe Streptococcus pneumoniae u dzieci do 5 roku zycia w wybranych srodowiskach w Warszawie. Pediatria Polska 78 | 2000 | Poland | 616 | Europe | CS | Unkn. | No | <5y | 1–5 | Unknown | 76 | 125 | 61% |
| | | 2001 | Poland | 616 | Europe | CS | Unkn. | No | <5y | 1–5 | Unknown | 63 | 120 | 52% |
| Sun (2007) | Epidemiological study of Streptococcus pneumoniae in the nasopharynx of healthy children under 5 years of age in Wuhan. (Chinese). Zhonghua Er Ke Za Zhi. Chinese Journal of Pediatrics 45 | 2003 | China | 156 | Asia | CS | Unkn. | No | <5y | 0–4 | Unknown | 135 | 605 | 22% |
| Syrjanen (2001) | Nasopharyngeal carriage of Streptococcus pneumoniae in Finnish children younger than 2 years old. Journal of Infectious Diseases 184 | 1994 | Finland | 246 | Europe | Long. | Rand. | No | <5y | 1–2 | Unknown | 649 | 3024 | 21% |
| Syrogiannopoulos (1997) | Resistance patterns of streptococcus pneumoniae from carriers attending day-care centers in southwestern Greece. Clinical Infectious Diseases 25 | 1995 | Greece | 300 | Europe | CS | Unkn. | No | <5y | 1–1 | Unknown | 9 | 20 | 45% |
| | | 1995 | Greece | 300 | Europe | CS | Unkn. | No | <5y | 2–4 | Unknown | 96 | 236 | 41% |
| | | 1995 | Greece | 300 | Europe | CS | Unkn. | No | 5–17y | 5–6 | Unknown | 27 | 82 | 33% |
| Syrogiannopoulos (2002) | Antimicrobial use and serotype distribution of nasopharyngeal Streptococcus pneumoniae isolates recovered from Greek children younger than 2 years old. Clinical Infectious Diseases 35 | 1997 | United States | 840 | Americas | CS | Outp. | No | <5y | 1–1 | Unknown | 395 | 1190 | 33% |
| | | 1997 | Greece | 300 | Europe | CS | Outp. | No | <5y | <1 | Unknown | 371 | 1258 | 29% |
| Syrogiannopoulos (2008) | Impact of heptavalent pneumococcal conjugate vaccine on nasopharyngeal carriage of penicillin-resistant Streptococcus pneumonide among day-care center attendees in central Greece. Pediatric Infectious Disease Journal 27 | 2005 | Greece | 300 | Europe | CS | Unkn. | No | <5y | 1–1 | Unknown | 8 | 16 | 50% |
| | | 2005 | Greece | 300 | Europe | CS | Unkn. | No | <5y | 2–4 | Unknown | 263 | 507 | 52% |
| | | 2005 | Greece | 300 | Europe | CS | Unkn. | No | 5–17y | 5–6 | 5 | 50 | 147 | 34% |
| Takala (1996) | Subtyping of common pediatric pneumococcal serotypes from invasive disease and pharyngeal carriage in Finland. Journal of Infectious Diseases 173 (1) | 1987 | Finland | 246 | Europe | CS | Outp. | No | <5y | Unreported | 3 | 50 | | Unkn. |
| Talarico (2010) | Epidemiologic characteristics of streptococcus pneumoniae in Vietnam and implications for population vaccination. Doctoral dissertation, University of Michigan | 2006 | Vietnam | 704 | Asia | CS | Rand. | No | <5y | 0–5 | Unknown | 42 | 104 | 40% |
| | | 2006 | Vietnam | 704 | Asia | CS | Rand. | No | 5–17y | 6–17 | Unknown | 6 | 55 | 11% |
| | | 2006 | Vietnam | 704 | Asia | CS | Rand. | No | 18+y | 18–86 | Unknown | 8 | 360 | 2% |
| | | 2013 | Cuba | 192 | Americas | CS | Rand. | No | <5y | <1.5 | Unknown | 48 | 195 | 25% |
| | | 2013 | Cuba | 192 | Americas | CS | Rand. | No | <5y | 0.5–1 | Unknown | 75 | 365 | 21% |

| | | | | | | | | | | | | | | |
|---|---|---|---|---|---|---|---|---|---|---|---|---|---|---|
| Toledo (2017) | Prevalence of Pneumococcal Nasopharyngeal Carriage Among Children 2-18 Months of Age: Baseline Study Pre Introduction of Pneumococcal Vaccination in Cuba. Pediatric Infectious Disease Journal 36 (1) | 2013 | Cuba | 192 | Americas | CS | Rand. | No | <5y | 1–1.5 | Unknown | 89 | 420 | 21% |
| Tóthpál (2012) | Nasal carriage of streptococcus pneumoniae among Hungarian children before the wide use of the conjugate vaccine. Acta Microbiologica et Immunologica Hungarica 59 (1) | 2009 | Hungary | 348 | Europe | CS | Unkn. | No | <5y | 3–6 | Unknown | 135 | 358 | 38% |
| Trojanek (2013) | Pneumococcal urinary antigen positivity in healthy colonized children: is it age dependent?. Wiener Klinische Wochenschrift 125 | 2010 | Czechia | 203 | Europe | CS | Unkn. | No | 5–17y | 3–6 | 5 | 105 | 197 | 53% |
| Trucco (1996) | Penicillin resistant Streptococcus pneumoniae among children attending day care centers. SpanishPrevalencia de Streptococcus pneumoniae resistente a penicilina en ninos que asisten a jardines infantiles en Santiago. Revista Chilena de Pediatria 67 | 1994 | Chile | 152 | Americas | CS | Unkn. | No | <5y | 1–4 | Unknown | 120 | 200 | 60% |
| Tsolia (1999) | Prevalence and patterns of resistance of Streptococcus pneumoniae strains isolated from carriers attending day care centers in the area of Athens. Microbial Drug Resistance 5 | 1997 | Greece | 300 | Europe | CS | Unkn. | No | <5y | 0–6 | 4 | 136 | 382 | 36% |
| Turner (2012) | A longitudinal study of Streptococcus pneumoniae carriage in a cohort of infants and their mothers on the Thailand-Myanmar border. PLOS ONE 7 | 2007 | Thailand | 764 | Asia | Long. | Outp. | No | <5y | <1 | 0 | 162 | 196 | 83% |
| | | 2007 | Thailand | 764 | Asia | Long. | Outp. | No | <5y | <1 | 0 | 149 | 184 | 81% |
| | | 2007 | Thailand | 764 | Asia | Long. | Outp. | No | <5y | 1–1 | 1 | 110 | 146 | 75% |
| | | 2007 | Thailand | 764 | Asia | Long. | Outp. | No | <5y | 1–1 | 1 | 119 | 154 | 77% |
| | | 2007 | Thailand | 764 | Asia | Long. | Outp. | No | 18+y | ≥18 | Unknown | 24 | 127 | 19% |
| | | 2007 | Thailand | 764 | Asia | Long. | Outp. | No | 18+y | ≥18 | Unknown | 50 | 179 | 28% |
| | | 2007 | Thailand | 764 | Asia | Long. | Outp. | No | 18+y | ≥18 | Unknown | 37 | 141 | 26% |
| | | 2007 | Thailand | 764 | Asia | Long. | Outp. | No | 18+y | ≥18 | Unknown | 45 | 192 | 23% |
| | | 2007 | Thailand | 764 | Asia | Long. | Outp. | No | 18+y | ≥18 | Unknown | 47 | 228 | 21% |
| Vakevainen (2010) | Serotype-specific hyporesponsiveness to pneumococcal conjugate vaccine in infants carrying pneumococcus at the time of vaccination. Journal of Pediatrics 157 | 2000 | Philippines | 608 | Asia | CS | Rand. | No | <5y | <1 | Unknown | 297 | 1059 | 28% |
| Veenhoven (2004) | Nasopharyngeal pneumococcal carriage after combined pneumococcal conjugate and polysaccharide vaccination in children with a history of recurrent acute otitis media. Clinical Infectious Diseases 39 | 1998 | Netherlands | 528 | Europe | CS | Rand. | No | <5y | 1–2 | 1 | 41 | | Unkn. |
| | | 1999 | Netherlands | 528 | Europe | CS | Rand. | No | <5y | 1–2 | 1 | 63 | | Unkn. |
| Verani (2018) | Nasopharyngeal carriage of Streptococcus pneumoniae leamong HIV-infected and - uninfected children <5 years of age before introduction of pneumococcal conjugate vaccine in Mozambique. PLOS ONE 13 (2) | 2012 | Mozambique | 508 | Africa | CS | Unkn. | No | <5y | 0–5 | Unknown | 255 | 320 | 80% |
| Wang (2003) | Rate of nasopharyngeal carriage, antimicrobial resistance and serotype of Streptococcus pneumoniae among children in northern Taiwan. Journal of Microbiology, Immunology and Infection 36 | 1998 | Taiwan | 158 | Asia | CS | Outp. | No | <5y | 0–1 | Unknown | 24 | 180 | 13% |
| | | 1998 | Taiwan | 158 | Asia | CS | Outp. | No | <5y | 2–5 | Unknown | 50 | 180 | 28% |
| | | 1998 | Taiwan | 158 | Asia | CS | Outp. | No | 5–17y | 6–14 | Unknown | 21 | 118 | 18% |
| Warda (2013) | Antibiotic resistance and serotype distribution of nasopharyngeal isolates of Streptococcus pneumoniae from children in Marrakech region (Morocco). Journal of Infection and Public Health 6 (6) | 2007 | Morocco | 504 | Africa | CS | Outp. | No | <5y | 0–2 | 1 | 302 | 660 | 46% |
| Wattal (2007) | Nasopharyngeal carriage of Streptococcus pneumoniae. Indian Journal of Pediatrics 74 (10) | 2000 | India | 356 | Asia | CS | Outp. | No | <5y | 1–3 | Unknown | 13 | 200 | 6% |
| Xie (2018) | A Cross-sectional Survey Assessing Carriage of Streptococcus pneumoniae in a Healthy Population in Xinjiang Uygur Autonomous Region of China. Biomedical and Environmental Sciences 31 (3) | 2015 | China | 156 | Asia | CS | Unkn. | Yes | 5–17y | 0–53 | 11 | 361 | 513 | 70% |
| Yagupsky (1998) | Acquisition, carriage, and transmission of pneumococci with decreased antibiotic susceptibility in young children attending a day care facility in southern Israel. Journal of Infectious Diseases 177 | 1993 | Israel | 376 | Asia | Long. | Unkn. | No | <5y | 1–3 | Unknown | 362 | 576 | 63% |
| Yeh (2003) | Heptavalent pneumococcal vaccine conjugated to outer membrane protein of Neisseria meningitidis serogroup b and nasopharyngeal carriage of Streptococcus pneumoniae in infants. Vaccine 21 | 1995 | United States | 840 | Americas | CS | Trial | No | <5y | 0–1 | 0 | 31 | | Unkn. |
| Yen-Chen, Yien-Lin (2011) | Nasopharyngeal carriage of Streptococcus pneumoniae in Taiwan before and after the introduction of a conjugate vaccine. Vaccine 29 | 2005 | Taiwan | 158 | Asia | CS | Outp. | No | <5y | 0–5 | Unknown | 195 | 1506 | 13% |
| | | 2006 | Taiwan | 158 | Asia | CS | Outp. | No | <5y | 0–5 | Unknown | 407 | 2633 | 15% |
| Yoshida (2011) | Association between nasopharyngeal load of Streptococcus pneumoniae, viral coinfection, and radiologically confirmed pneumonia in Vietnamese children. Pediatric Infectious Disease Journal 30 | 2008 | Vietnam | 704 | Asia | CS | Rand. | No | <5y | 0–4 | Unknown | 175 | 350 | 50% |

| | | | | | | | | | | | | | | |
|---|---|---|---|---|---|---|---|---|---|---|---|---|---|---|
| Zemlickova (2006) | Characteristics of Streptococcus pneumoniae, Haemophilus influenzae, Moraxella catarrhalis and Staphylococcus aureus isolated from the nasopharynx of healthy children attending day-care centres in the Czech Republic. Epidemiology and Infection 134 | 2004 | Czechia | 203 | Europe | CS | Unkn. | No | <5y | 3–6 | Unknown | 162 | 425 | 38% |
| Zemlickova (2010) | Serotype-specific invasive disease potential of Streptococcus pneumoniae in Czech children. Journal of Medical Microbiology 59 (Pt 9) | 2004 | Czechia | 203 | Europe | CS | Unkn. | No | <5y | 0–5 | Unknown | 138 | | Unkn. |
