## Supplementary files for "Global landscape of *Streptococcus pneumoniae* serotypes colonising healthy individuals worldwide before vaccine introduction; a systematic review and meta-analysis"

### Appendices

*Appendix 1: Search terms and screening strategy*
All references were compiled in an EndNote database.

*Search terms*
The search terms used were based on a combination of the ‘Pathogen’ search and the ‘Colonization’ search, with no restrictions.
(“streptococcus pneumoniae” ([MeSH] if MEDLINE, database-specific subject headings for other databases) OR “pneumococcus” [all fields] OR “pneumococcal” [all fields] OR “s. pneumoniae” [all fields] OR “streptococcal” [all fields] OR “streptococcus” [all fields])
AND
(“colonization” [all fields] OR “colonisation” [all fields] OR “carriage” [all fields] OR “carrier” [all fields] OR “colonised” [all fields]  OR “colonized” [all fields])

The search was conducted on EMBASE, MEDLINE, The Cochrane Library, Web of Science, Biological Abstracts (BIOSIS citation index), Global Health (including the Public Health and Tropical Medicine (PHTM) database and the human health and diseases information extracted from CAB Health), Cumulative Index to Nursing and Allied Health Literature (CINAHL), Psychological abstracts (PsycINFO), Latin America and Caribbean Health Sciences Info (LILACS), and Africa-Wide Info.

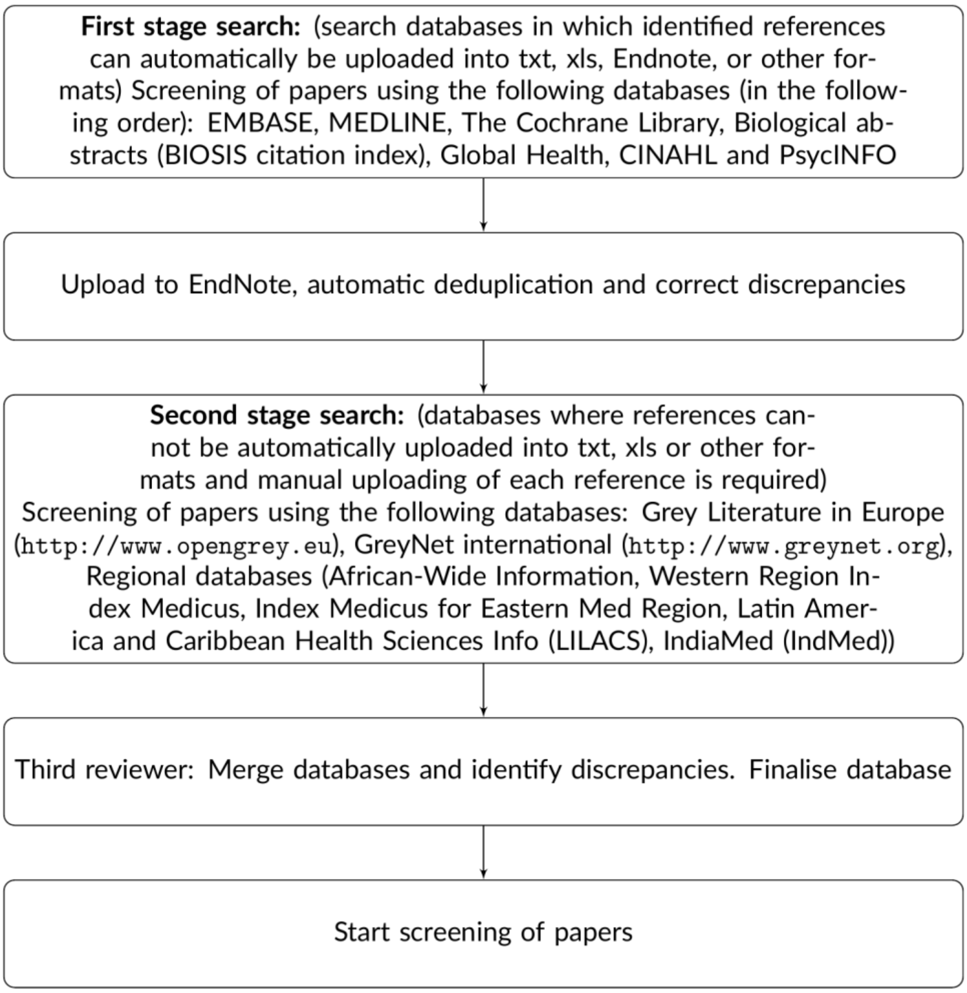


Figure S1: Flowchart of search strategy

#### *Screening of articles*

##### *Inclusion criteria*

(i) Study on *Streptococcus pneumoniae*
(ii) Providing carriage estimates from nose and/or throat
(iii) Not inpatients
(iv) Individuals not vaccinated with PCV
(v) Study participants not selected based on the presence or absence of pneumococcal-like illness (including acute respiratory infection, sinusitis, acute otitis media, sepsis, meningitis, and pneumonia)
(vi) Settings in which PCV had not yet been introduced, including control arms of cluster-randomised intervention studies and individual-based intervention studies in which <20% of the study population in the targeted age group had received PCV.

We restricted the study to primary studies; when a number of publications emanated from the same dataset, we pooled those studies under one umbrella study. Conference abstracts were only included if they had not yet been published as full articles.

Studies were only included if they were primary or co-primary (one of many articles reporting a subset of the study data).

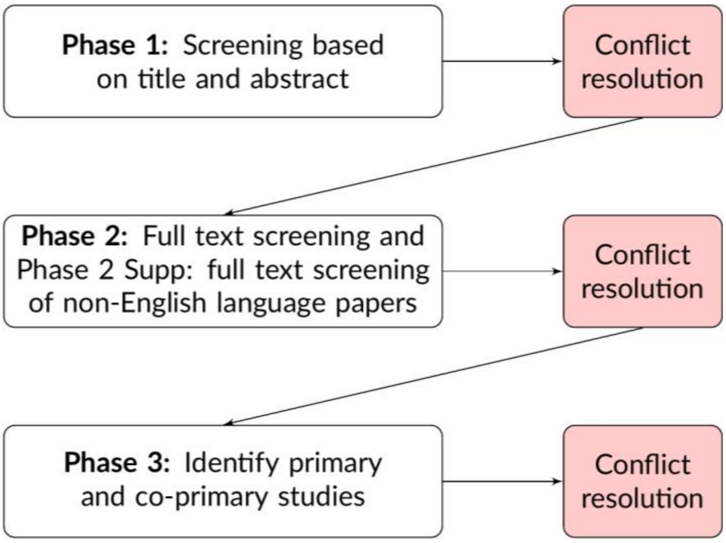


Figure S2: Flowchart of abstract and full text screening. Primary studies are those which report a full carriage survey; co-primary studies are those publications which report a subset of the collected data.

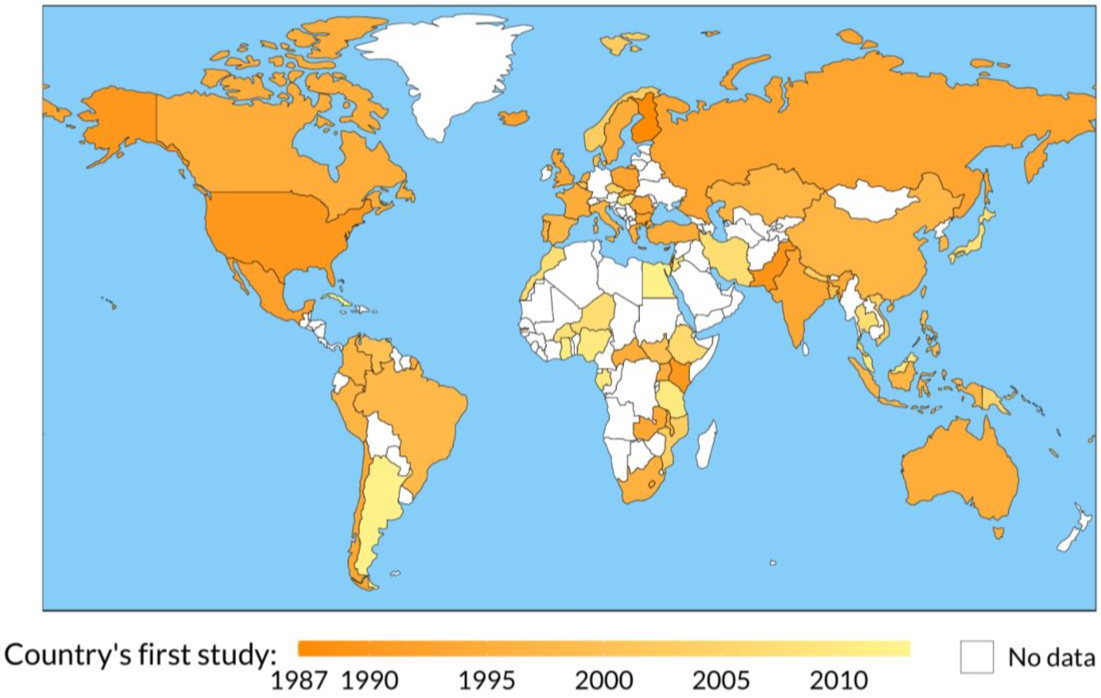


Figure S3: Starting year of first serotype carriage survey conducted in each country. The earliest studies were in Finland (1987), Pakistan (1989), and USA, Kenya and The Gambia (1990). Map shapefile from Natural Earth (public domain).

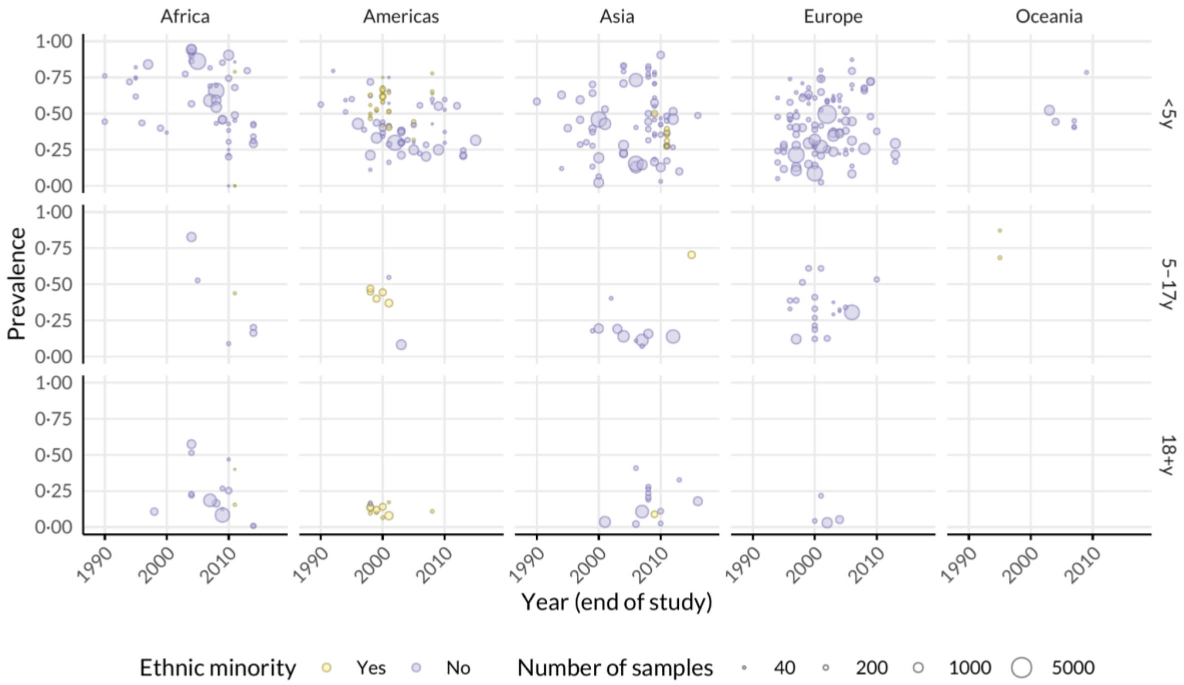


Figure S4: Prevalence of carriage in included studies, stratified by age group, continent, and ethnic minority status.

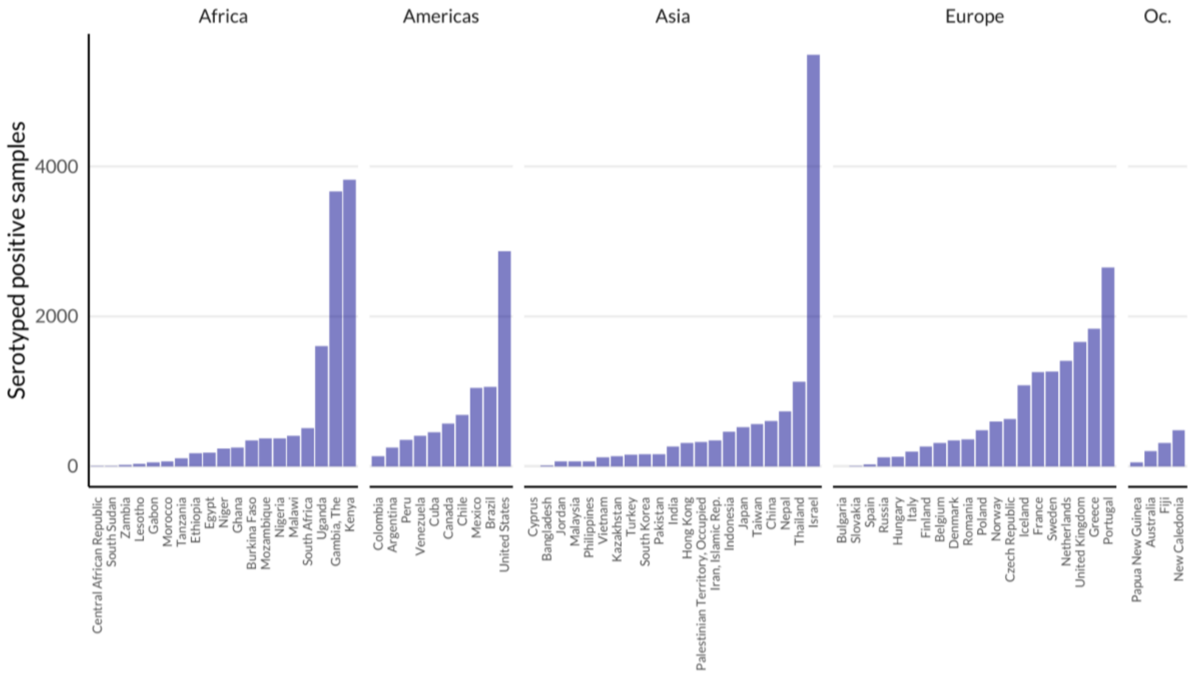


Figure S5: Number of serotyped positive samples in each country, grouped by continent (NB: Oceania is represented as Oc.).

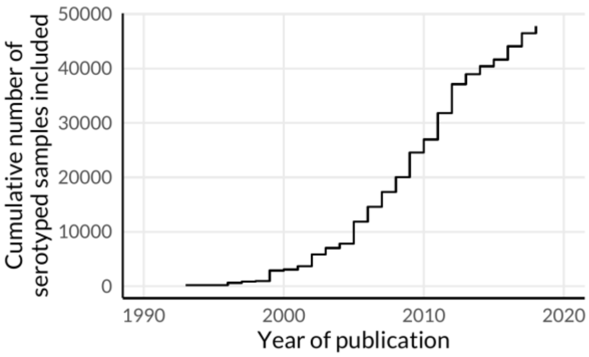


Figure S6: Cumulative number of serotyped pneumococcal carriage isolates reported by studies in pre-PCV settings.

*Appendix 2: Analysis*

*Model structure*

Our main study outcome was the serotype distribution of *Streptococcus pneumoniae* in naso/oropharyngeal (NP/OP) carriage. We estimated such an outcome by age group and geographical area (country, subcontinent, and continent). We considered the three age groups: <5 years, 5–17 years, and 18+ years.

In the simplest approach we denote $N_k = (N_{k,1}, \dots, N_{k,G})$ as the row vector of the observed number of individuals in dataset $k$ for each of the $G$ serogroups considered. We assume that the data follow a multinomial sampling distribution with true frequencies in each serogroup of $\Pi_k = (\Pi_{k,1}, \dots, \Pi_{k,G})$, and the sum of probabilities $\sum_{g=1}^{G} \Pi_{k,g} = 1$. Within each dataset, in each serogroup, $g$, there is a distribution of individuals in each serotype $N_{k,g} = \left(N_{k,g,1}, \dots, N_{k,g,S_g}\right)$ where $S_g$is the number of serotypes in serogroup $g$, such that there are $T = \sum_{g=1}^{G} S_g$ serotypes overall. This serotype data are also assumed to follow a multinomial sampling distribution, from some true frequencies, $\Pi_{k,g} = \left(\Pi_{k,g,1}, \dots, \Pi_{k,g,S_g}\right)$, where $\sum_{s=1}^{S_g} \Pi_{k,g,s} = 1$, and is the conditional probability (or true frequency) of being in serotype $s$ given serogroup $g$. The vector of probability $\Pi_{k,g}$ is then calculated for each serogroup containing multiple serotypes, and is equal to 1 if the serogroup is only a single serotype, e.g. serotype 4 is the complete serogroup 4. The serotype distribution can thus be estimated as the product of the serogroup distribution by that of the serotype distribution within each serogroup, such that $\Pi_{k,s} = \Pi_{k,g}\Pi_{k,g,s}$is the probability distribution of serotypes $s$ in dataset $k$.

In the meta-analysis, the probabilities were then pooled by geographical level and age group. Thus, for setting, $x$, (i.e. continent and age group) we computed one vector of probabilities for serogroups $\Pi_x$ and vectors of probabilities for the serotypes in each of the serogroups, $\Pi_{x,g}$, and one vector for all the individual serotypes, $\Pi_{x,s}$, using information from all the datasets within $x$.

Priors for the vectors of probabilities followed a Dirichlet distribution with parameters $\alpha_k = 1, \alpha_{S_g} = 1$ for all serogroups and serotypes. Posterior distributions were sampled 10,000 times using Markov Chain Monte Carlo (MCMC) iteration in the rjags package in R.[51]

For the overall carriage model, we assumed each dataset was Binomial with probability of carriage unique to each age group and continent, $p_{a,c}$. The logit of $p_{a,c}$, $\beta_{a,c}$, had a hierarchical prior, $\beta_{a,c} \sim N(\gamma_a, \tau_\beta)$, with the age group level parameters having prior, $\gamma_a \sim N(0, \tau_\gamma)$. These normal distributions are parameterised with their precisions, $\tau = \sigma^{-2}$, and the standard deviations are given PC priors[52], $\sigma \sim \Gamma(1, 1.5)$.,

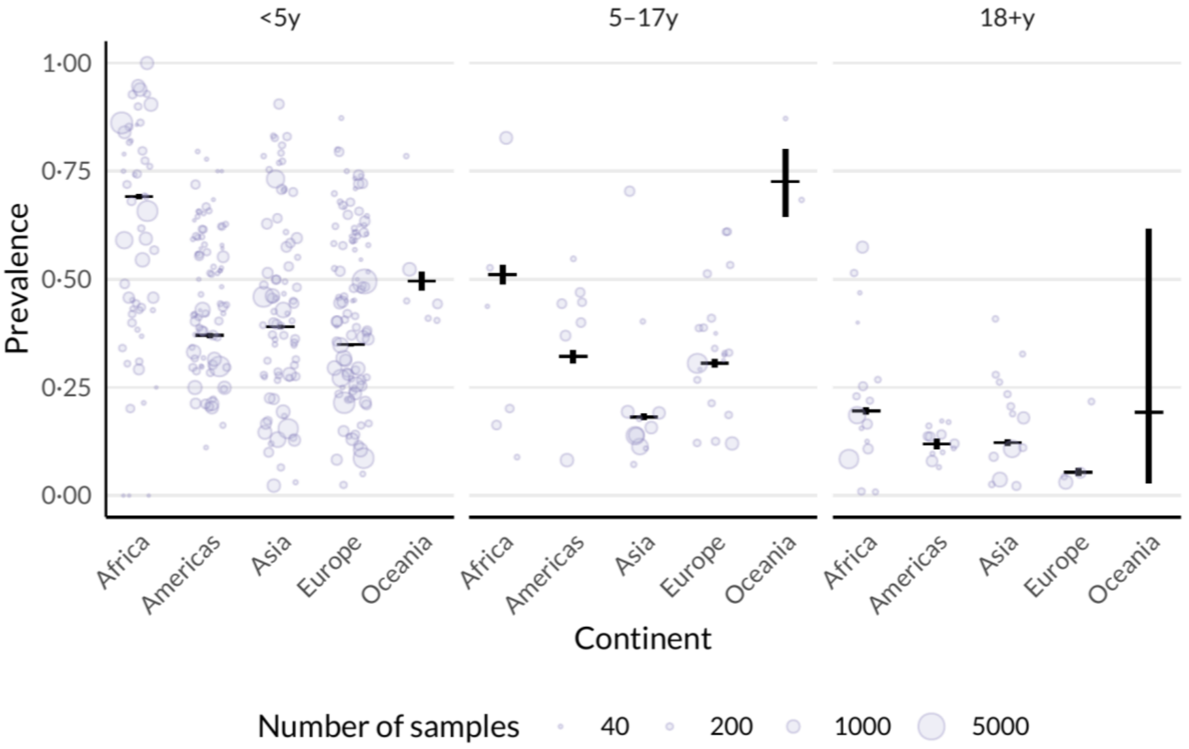


Figure S7: Modelled age-stratified regional carriage prevalence. Horizontal ticks indicate median prevalence, and error bars indicate 95% posterior quantiles. No study reported adult carriage in Oceania.

We considered the proportion of carriage events in each continent and age group which would be covered by a variety of PCV formulations (Table S1).

Table S1: Serotypes included in vaccine formulations considered for coverage.

| **Product** | 1 | 5 | 6B | 7F | 9V | 14 | 19F | 23F | 4 | 18C | 6A | 19A | 3 | 22F | 33F | 8 | 10A | 11A | 12F | 15B |
|---|---|---|---|---|---|---|---|---|---|---|---|---|---|---|---|---|---|---|---|---|
| Synflorix-10 | x | x | x | x | x | x | x | x | x | x | | | | | | | | | | |
| Pneumosil-10 | x | x | x | x | x | x | x | x | | | x | x | | | | | | | | |
| Prevnar-13 | x | x | x | x | x | x | x | x | x | x | x | x | x | | | | | | | |
| Vaxneuvance-15 | x | x | x | x | x | x | x | x | x | x | x | x | x | x | x | | | | | |
| Prevnar-20 | x | x | x | x | x | x | x | x | x | x | x | x | x | x | x | x | x | x | x | x |

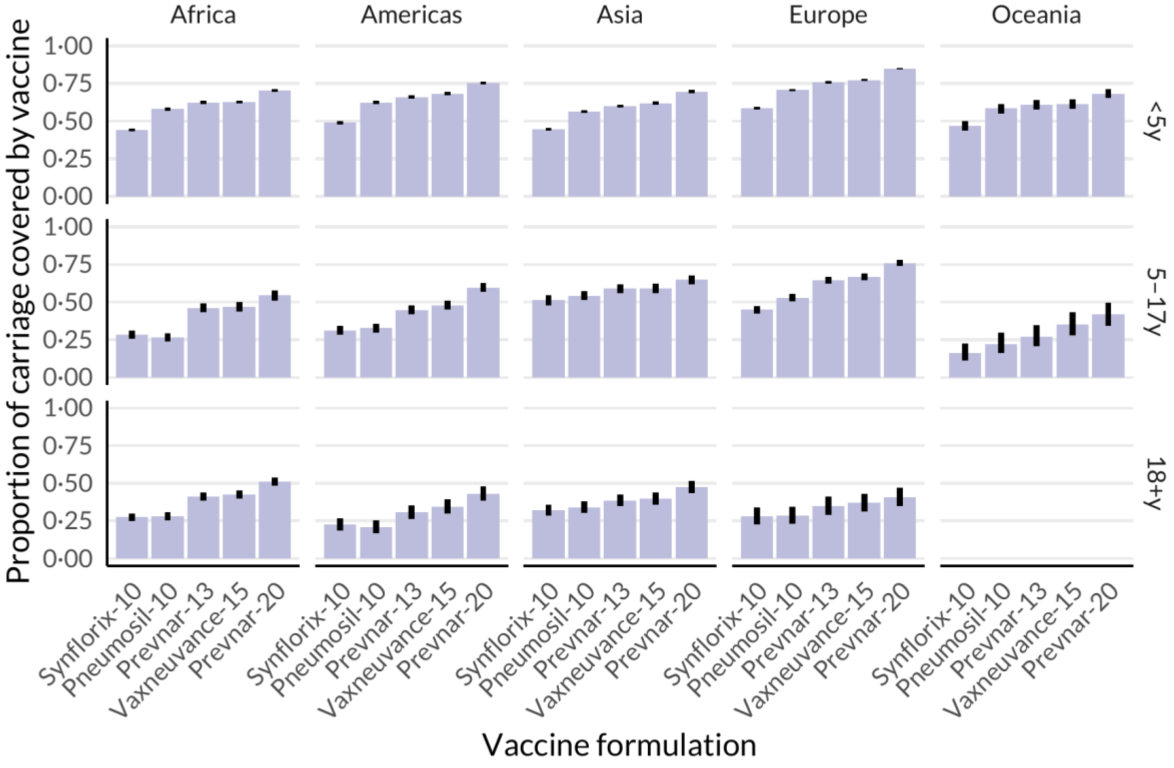


Figure S8: Proportion of carriage covered by the formulation in each vaccine product, stratified by continent and age group. Bars represent median estimates and error bars are 95% credible intervals. As in Figure 3 but with vaccine product and age swapped.

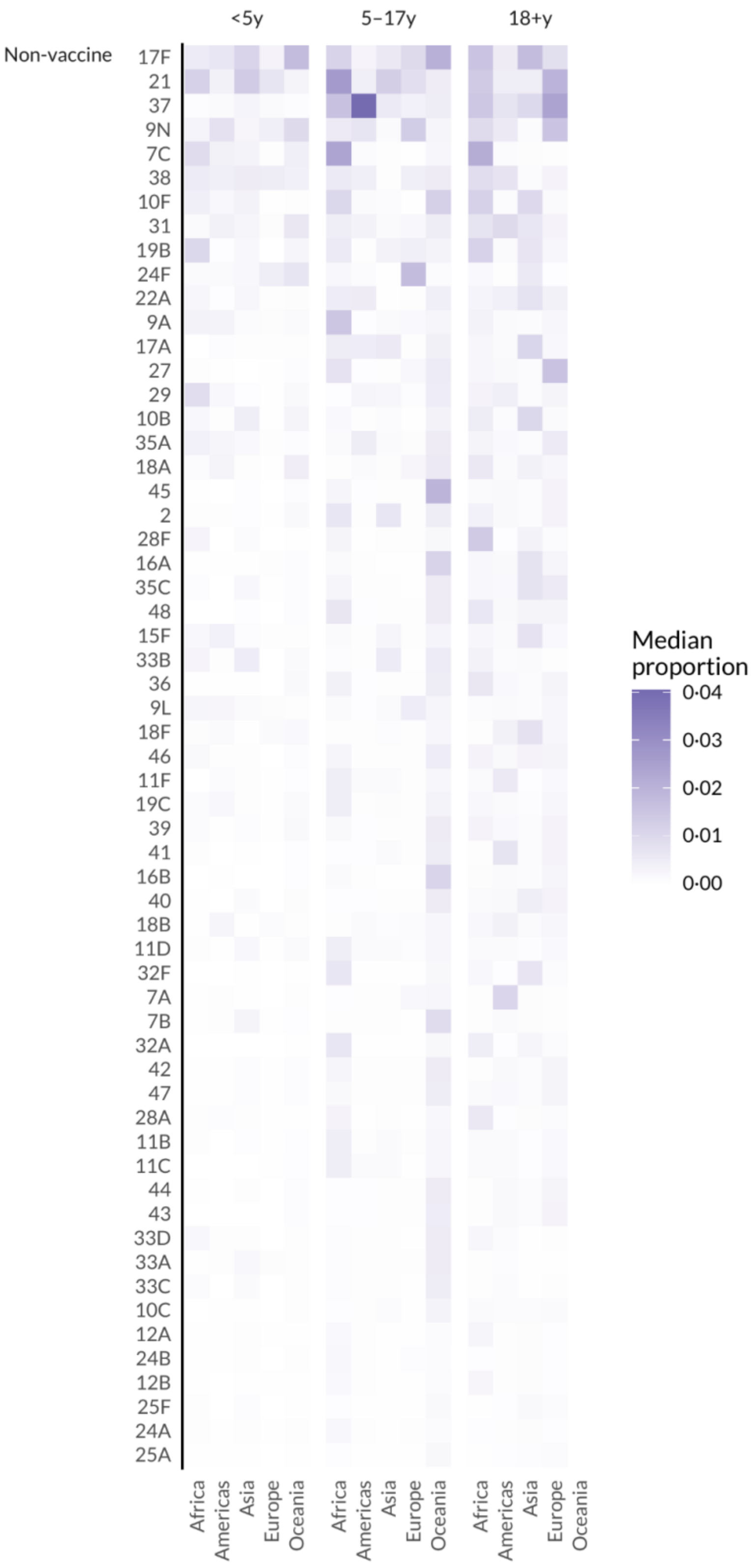


Figure S9: Estimated serotype prevalence by region and age category. Serotype ordering is by global median prevalence, within groupings of: serotypes in PCV10, additional serotypes in PCV13 vaccine, all non-vaccine serotypes, and the non-serotyped isolates. Numerical values are provided in Appendix 6.

*Population weighting*

Global ranking of median serotype prevalence was performed for each age group by calculating the average of each serotype's prevalence, weighted by each continent's mean population in that age group for the years between 1990 and 2015. Data were sourced from the United Nations' World Population Prospects 2019[53] whose age categories do not map directly to ours. Hence the 5–17y group contains the 5–9, 10–14 and 15–19 year olds, and the 18+y group is all ages 20 or older.

*Clusters*

For two discrete valued probability densities, $p, q$, over the $T$ serotypes, such that $p(x_i) = p_i$ and $q(x_i) = q_i$ are, respectively, the relative prevalence of serotype $i$ in each distribution,, the Bhattacharyya distance is calculated as

$$d(p(x), q(x)) = -\log\left(\sum_{i=1}^{T} \sqrt{p_i\, q_i}\right)$$

and takes on a value $d \geq 0$. This distance measure is used to calculate the similarity between studies for the clustering. For pairs which had no serotypes in common, the infinite distance between them was replaced with a value twice the maximum observed finite difference to ensure the clustering was computable. Distance-based clustering was chosen over principal component analysis (PCA) or other techniques due to each observation being composed of a vector of proportions adding to one, rather than a collection of correlated but independently measured variables.

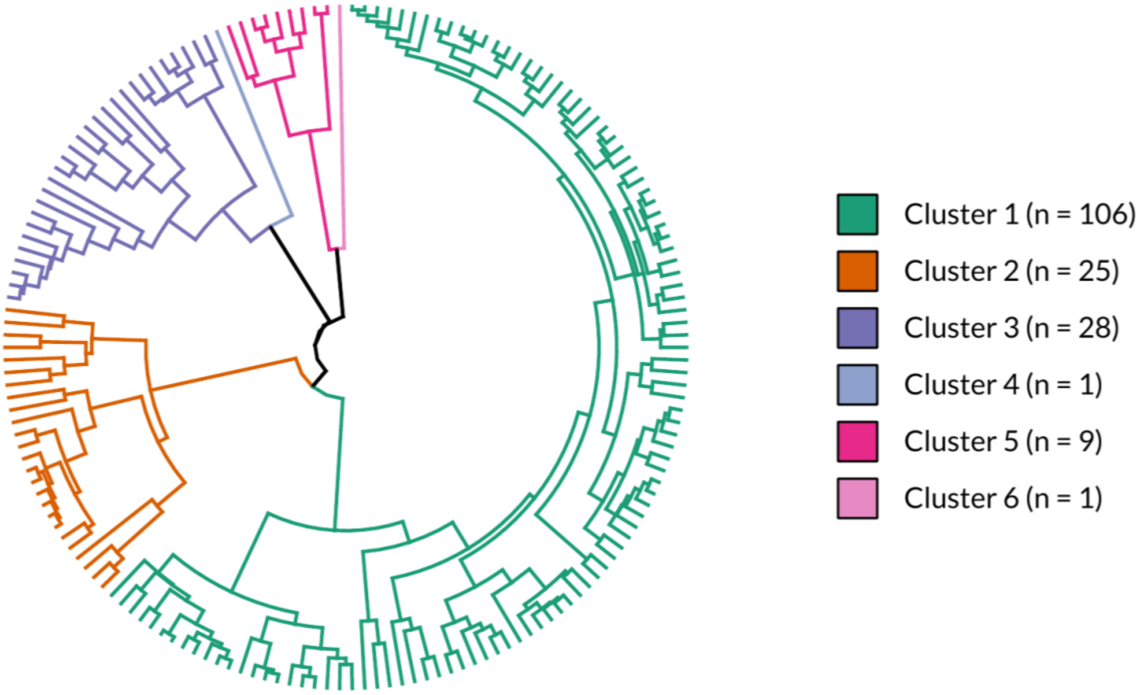


Figure S10: Dendrogram showing the clustering of serotype distributions and number of studies in each cluster. The radial axis has been truncated to 0·25 the maximum observed Bhattacharyya distance, which exaggerates how close the three branches composed of clusters 1/2, 3/4, and 5/6 are.

The optimal number of clusters according to the elbow method (largest increase in explained variation) was four, using a measure of total Bhattacharyya distance. Further inspection of the dendrogram (Figure S10) indicated that there may be two singleton clusters where each of the contained studies was markedly different to the remainder of the larger cluster from which they came.

Cluster 1 (n=106) contains most of the datasets on young children (92/132 (70%)) and contains a higher proportion of young childrens' datasets (92/106 (87%)) than if there was no clustering (132/170 (78%)). It also contains most of the datasets from Europe (49/56 (88%)) and while it had a lower proportion of datasets on ethnic minority groups (13/106 (12%)) than if there was no clustering (34/171 (20%)), nearly half the datasets on Native Americans (13/30 (43%)) were found in this cluster, and all 13 datasets were on under 5s.

Cluster 2 (n=25) contained most of the African studies (14/20 (70%)), had more adult datasets (4/25 (16%)) than if there was no clustering (14/170 (8%)) and a low proportion of ethnic minority datasets (3/25 (12%)).

A majority of the datasets in cluster 3 (n=28) are American (16/28 (57%)) and this cluster has the highest proportion of datasets drawn from ethnic minority populations (14/28 (50%)) compared to the overall data (20%). This cluster contains all datasets for Native American groups aged 5 years and over (10 datasets), with young Native American children being found here (4 datasets) and in clusters 1 (13), 2 (2) and 5 (1).

Cluster 4 (n=1) is most similar to cluster 3 and consists of a dataset of 32 5–17 year-old indigenous Babongo people in Gabon, with 44% carriage, dominated by serotypes 15A (38%), 3 (23%), and 11A (15%) (serotypes 34, 17F and 14 each represent 8%). The other remaining data from Gabon is from the same study of Babongo people and is either in cluster 2 (21 young children with 15 serotyped isolates) or was excluded for low power (50 adults with only 9 serotyped isolates).

Cluster 5 (n=9) contains both datasets on young indigenous Venezuelans and one on young Native Americans. The remainder of the cluster is four datasets on young Asian children under 5, and one dataset each on non-minority Brazilian children and Chilean adults.

Cluster 6 (n=1) is most similar to cluster 5. It consists of one dataset from a study on 142 general population Israeli adults with high prevalence (41%) and carriage dominated by serotype 5 (60%). All other Israeli datasets in the clustering are from the same study on young children (distinct from this one in cluster 6) and are found in clusters 1 (7) and 5 (1).

Clusters did not appear to differ greatly in terms of when the datasets' studies commenced. Apart from the singleton clusters, 4 (2005) and 6 (201), clustered datasets were from: 1995–2011 (Cluster 1), 1998–2012 (Cluster 2), 1998–2008 (Cluster 3), or 1996–2008 (Cluster 5).

$\chi^2$ tests of independence indicate that there are differences in age category, continent and ethnic minority status across clusters ($p$ < 0·001) but not relative prevalence ($p$ > 0·8). Training a *k*-nearest neighbours (kNN[54]) algorithm on the four clusters (1, 2, 3+4, 5+6) yields a classifier with low balanced accuracy according to Bhattacharyya distance (0·29). When training a kNN using the setting features, accuracy is 0·58 in the training and 0·46 in testing using leave-one-out cross-validation. This indicates that clustering studies and partitioning the variable space does not yield a way of accurately predicting the carried serotype distribution knowing the target age category, continent, whether carriage is high or low in terms of global carriage and whether the target population is an ethnic minority group.

Figure S11 shows the difference between ethnic minority and general population datasets across the three age groups in datasets used for the cluster analysis.

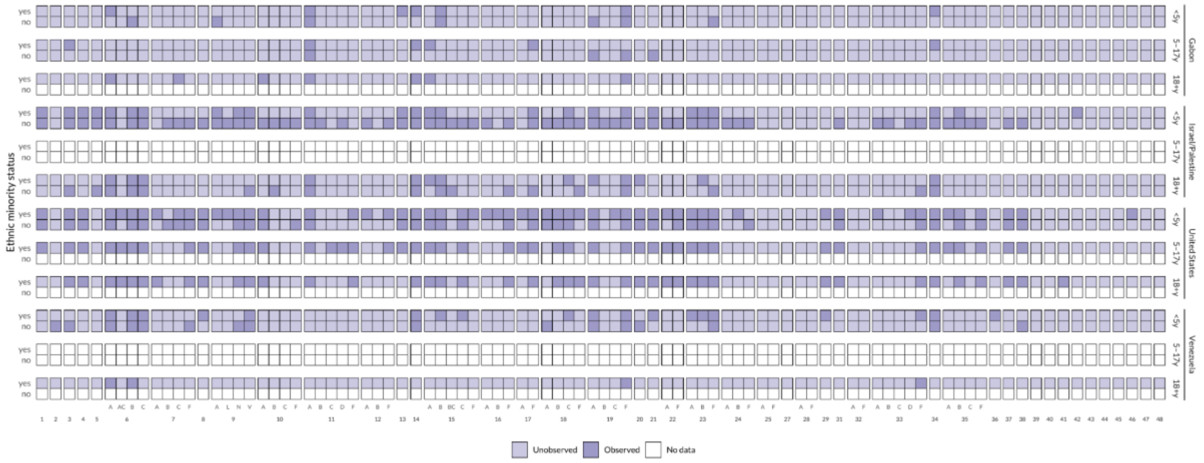


Figure S11: Serotypes observed in the general population and in ethnic minority groups in countries where more than 10 serotyped isolates sampled from ethnic minority populations were available.

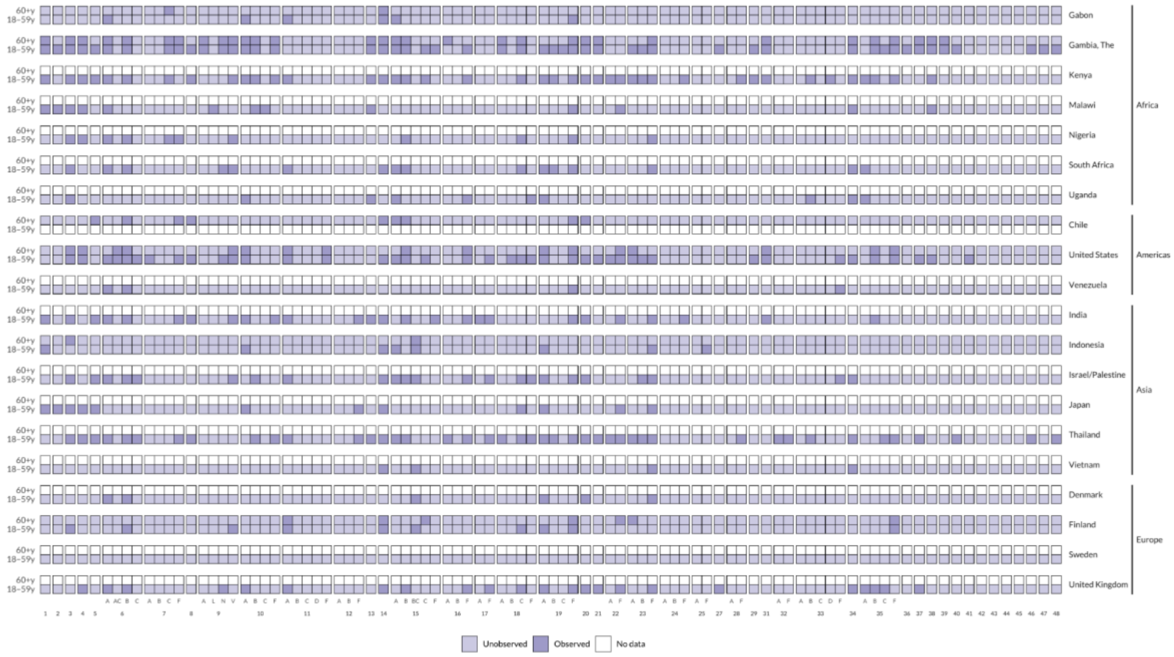


Figure S12: Serotypes observed in the elderly (aged 60 or more years) and non-elderly (aged 18–59 years) adult population for countries where adult carriage has been studied.

#### *Serotype diversity*

Simpson's index of diversity, $\lambda$, was calculated from the MCMC samples of $\Pi$ as a measure of pneumococcal diversity for serotype distribution[38,39] and is reported as the Gini-Simpson index

$(1 - \lambda)$ to describe the probability of randomly drawn samples belonging to different serotypes, and as the Inverse Simpson index $(\lambda^{-1})$ to describe the effective number of groups of co-occurring serotypes.

The Gini coefficient[55] was also calculated to evaluate the diversity of the serotype distribution, as twice the difference between the modelled cumulative prevalence distribution and that of a uniform random variable. It takes on values between 0 (flat serotype distribution) and 1 (all modelled carriage attributable to a single serotype).

*Data handling, assumptions and possible confounding*

Data were collected on studies with different designs, endpoints and sampling methodologies. Hence, a series of assumptions were made for the analysis; the robustness of some of those were explored through sensitivity analysis.

*Longitudinal studies:* In longitudinal studies, in which individuals are being swabbed multiple times, we averaged out the numerator and denominator over the study period, within each particular pre-defined age group. In doing so, we therefore assumed stability in serotype distribution over the age group and study period and did not take lack of independence (i.e. same individuals swabbed) into account.

*Changes over time:* It is likely that the serotype distribution in carriage varies over time in the same geographic setting, due to long secular trends as well as outbreaks. In our analysis, we pooled data over the years between 1990 and 2015, in line with the global serotype distribution in the IPD project.

*Age range*: Given that the age range of reported nasopharyngeal and oropharyngeal carriage distribution differs across studies, we allowed for some flexibility around exact age bands and explored through sensitivity analysis how results differ when the age cut-offs for inclusion are being widened. For <5y olds, we considered a difference of 1 year to be compared to the upper cut-off to be compatible (e.g. a number of studies provided data for children <6y olds rather than <5yr olds).

*Lack of serotype-specific differentiation*: For cross-reactive serotypes such as 6A/C and 15B/C, we initially analysed those serotypes as grouped, and reallocated the proportion from the combined cross-reactive serotype value to the constituent serotypes proportional to their relative abundance after fitting the model.

*Multiple serotypes:* Multiple serotypes per individual are infrequently reported, as assessed during a preliminary analysis of extracted data in RESPICAR. When multiple serotypes were reported, equal weights were given to the serotypes reported and the total number of isolates were considered as the denominator for the analysis of serotype distribution.

*Serotyping methods:* For each study we collected specific information about their laboratory methods. In our analysis we made the assumption that data from various laboratory methods were indeed comparable.

*Epidemiological sampling design:* We explored the impact of the study design on estimates obtained, particularly differences between studies in which participants have been sampled from the wider community (random/quasi-random sampling) to studies in which participants have been sampled from specific groups or clusters, such as schools, day-care centres of workplaces.

*Appendix 3: Diversity results*

Table S2: Gini coefficients for modelled serotype distributions (medians and 95% intervals). A value of 1 indicates all carriage is attributed to one serotype; a value of 0 indicates uniformity.

| Continent | <5y | 5–17y | 18+y |
|---|---|---|---|
| Americas | 0·82 (0·81, 0·82) | 0·75 (0·73, 0·76) | 0·70 (0·68, 0·73) |
| Asia | 0·78 (0·77, 0·78) | 0·84 (0·83, 0·85) | 0·68 (0·65, 0·71) |
| Africa | 0·79 (0·78, 0·80) | 0·66 (0·64, 0·68) | 0·65 (0·63, 0·67) |
| Europe | 0·86 (0·86, 0·87) | 0·81 (0·80, 0·82) | 0·72 (0·68, 0·76) |
| Oceania | 0·79 (0·77, 0·81) | 0·65 (0·59, 0·70) | ·· |

Table S3: Inverse Simpson indices, $1/\lambda$, for modelled serotype distributions (medians and 95% intervals); interpretable as the effective number of groups of co-occurring serotypes circulating.

| Continent | <5y | 5–17y | 18+y |
|---|---|---|---|
| Americas | 15 (15, 16) | 28 (25, 31) | 31 (29, 33) |
| Asia | 13 (13, 14) | 23 (21, 24) | 24 (21, 28) |
| Africa | 16 (15, 16) | 13 (12, 14) | 22 (19, 25) |
| Europe | 11 (11, 11) | 16 (15, 18) | 15 (12, 20) |
| Oceania | 15 (14, 16) | 30 (22, 37) | ·· |

Table S4: Gini-Simpson indices, $1 - \lambda$, for modelled serotype distributions (medians and 95% intervals); interpretable as the probability that two isolates in the same setting are the same serotype.

| Continent | <5y | 5–17y | 18+y |
|---|---|---|---|
| Americas | 0·94 (0·93, 0·94) | 0·96 (0·96, 0·97) | 0·97 (0·97, 0·97) |
| Asia | 0·93 (0·92, 0·93) | 0·96 (0·95, 0·96) | 0·96 (0·95, 0·96) |
| Africa | 0·94 (0·93, 0·94) | 0·92 (0·92, 0·93) | 0·95 (0·95, 0·96) |
| Europe | 0·91 (0·91, 0·91) | 0·94 (0·94, 0·94) | 0·94 (0·91, 0·95) |

| | | | |
|---|---|---|---|
| Oceania | 0·93 (0·93, 0·94) | 0·97 (0·95, 0·97) | ·· |

**Appendix 4: Data entry forms**
Provided as a separate document.

*Appendix 5: Detailed characteristics of the studies*
Provided as a separate document.

A library of the cited works is available on Zotero.
